# Interim results from comparison of immune responses elicited by an inactivated and a vectored SARS-CoV-2 vaccine in seronegative and seropositive participants in India

**DOI:** 10.1101/2023.01.03.22284082

**Authors:** Mangaiarkarasi Asokan, Roshni Florina Joan, Sudhir Babji, Girish Dayma, Prajitha Nitheesh, Vinutha Subrahmanyam, Archana Pandey, Girish Malagi, Pooja Arya, Vibhuti Mahajan, Jayateerth Bhavikatti, Ketakee Pawar, Aishwarya Thorat, Priyanki Shah, Ramakrishna B Goud, Bishnudeo Roy, Shon Rajukutty, Sushil Immanuel, Dhiraj Agarwal, Sankhanil Saha, Akshatha Shivaraj, Patricia Panikulam, Rajeshwari Shome, Shah-E-Jahan Gulzar, Anusmrithi U Sharma, Ajinkya Naik, Shruti Talashi, Madhuri Belekar, Ritu Yadav, Poornima Khude, V Mamatha, S Sudarshan, Urmila Deshmukh, Chinmayee Bhise, Manjiri Joshi, Leeberk Raja Inbaraj, Sindhulina Chandrasingh, Aurnab Ghose, Colin Jamora, Anandi S Karumbati, Varadharajan Sundaramurthy, Avita Johnson, Naveen Ramesh, Nirutha Chetan, Chaitra Parthiban, Asma Ahmed, Srabanti Rakshit, Vasista Adiga, George D’souza, Vinay Rale, Carolin Elizabeth George, Jacob John, Anand Kawade, Akanksha Chaturvedi, Anu Raghunathan, Mary Dias, Anand Bhosale, Padinjat Raghu, LS Shashidhara, Annapurna Vyakarnam, Vineeta Bal, Gagandeep Kang, Satyajit Mayor

## Abstract

**Background:** There are limited global data on head-to-head comparisons of vaccine platforms assessing both humoral and cellular immune responses, stratified by pre-vaccination serostatus. The COVID-19 vaccination drive for the Indian population in the 18 to 45-year age-group began in April 2021 when seropositivity rates in the general population were rising due to the Delta wave in April-May 2021.

**Methods:** Between 30 June 2021 and 28 January 2022, we enrolled 691 participants in the 18-45 age group across 4 clinical sites in India. In this non-randomized and laboratory blinded study, participants received either two doses of Covaxin^®^ 4 weeks apart or two doses of Covishield™ 12 weeks apart per the national vaccination policy. The primary outcome was the seroconversion rate and the geometric mean titer (GMT) of antibodies against the SARS-CoV-2 spike and nucleocapsid proteins. The secondary outcome was the frequency of cellular immune responses pre- and post-vaccination.

**Findings:** When compared to pre-vaccination baseline, both vaccines elicited statistically significant seroconversion and binding antibody levels in both seronegative and seropositive individuals. In the per-protocol cohort, Covishield™ elicited higher antibody responses than Covaxin^®^ as measured by seroconversion rate (98.3% vs 74.4%, p<0.0001 in seronegative individuals; 91.7% vs 66.9%, p<0.0001 in seropositive individuals) as well as by anti-spike antibody levels against the ancestral strain (GMT 1272.1 vs 75.4 BAU/ml, p<0.0001 in seronegative individuals; 2089.07 vs 585.7 BAU/ml, p<0.0001 in seropositive individuals). Not all sites recruited at the same time, therefore site-specific immunogenicity was impacted by the timing of vaccination relative to the Delta and Omicron waves. Surrogate neutralizing antibody responses against variants-of-concern were higher in Covishield™ recipients than in Covaxin^®^ recipients and in seropositive than in seronegative individuals after both vaccination and asymptomatic Omicron infection. T cell responses are reported from only one of the four site cohorts where the vaccination schedule preceded the Omicron wave. In seronegative individuals, Covishield™ elicited both CD4+ and CD8+ spike-specific cytokine-producing T cells whereas Covaxin^®^ elicited mainly CD4+ spike-specific T cells. Neither vaccine showed significant post-vaccination expansion of spike-specific T cells in seropositive individuals.

**Interpretation:** Covishield™ elicited immune responses of higher magnitude and breadth than Covaxin^®^ in both seronegative individuals and seropositive individuals, across cohorts representing the pre-vaccination immune history of the majority of the vaccinated Indian population.

## Introduction

As of 14^th^ December 2021, inactivated, vectored and mRNA vaccines comprised 43, 28 and 26% respectively of the total 10.6 billion doses of Covid vaccines administered in the world^1^. Despite this large share of the vaccination landscape, there are few studies that make comprehensive head-to-head comparisons of both humoral and cellular immune response elicited by these vaccine platforms^2–5^. As of 1^st^ September 2022, India has administered 1.7 billion doses of Covishield™ (ChAdOx-1 nCoV-19, manufactured by Serum Institute of India) and 0.35 billion doses of Covaxin^®^ (inactivated vaccine adjuvanted with a TLR7/8 agonist, manufactured by Bharat Biotech)^6^. These two vaccines administered in India, together represent 15% of the total Covid vaccine doses administered in the world. However, there is a paucity of data comparing the immunogenicity of the two vaccines.

Prior studies comparing Covishield™ and Covaxin^®^ were limited to addressing only antibody responses and in particular in the health care worker population who were immunized prior to the delta wave^7–11^ (See Supplementary Table 1). There is however, limited data on cellular immune responses elicited by these vaccines and no direct head-to-head comparisons or stratification by pre-vaccination serostatus^11^.

Vaccination following exposure to the Delta or Omicron variants of SARS-CoV-2 is likely to affect the quality, quantity and duration of immune responses. For determining future COVID-19 vaccination policy when pan-coronavirus or sarbecovirus vaccines become available, it becomes important to take into account pre-vaccination immune history of the majority of the vaccinated Indian population. Comparison of vaccine platforms stratified by serostatus will also inform vaccine development against future pandemics that are increasingly likely with climate change, ecosystem destabilization and lowering herd immunity due to vaccine hesitancy against both established and newer vaccine regimes.

## Results

### Study population

Between 30 June 2021 and 28 January 2022, we enrolled 691 participants in the 18-45 age group across 4 sites in urban and rural Bengaluru and Pune, India. The 4 clinical sites recruited participants in a staggered fashion and coincide with the different waves of vaccination in India following the Delta wave when vaccines became widely available (Figure 1). As per the prevailing government norms, participants received either two doses of Covaxin^®^ at 28 days apart or two doses of Covishield™ at 3 months apart. The Omicron wave in early 2022 overlapped with the second dose of vaccine in 2 sites and with both doses in 1 site. Participants were sampled at 6 timepoints for antibody analyses and at 4 timepoints for cellular analyses. The vaccine groups were subdivided as seronegative or positive at baseline based on binding antibody titers against the trimeric spike protein using a globally comparable and standardized test, the Diasorin TrimericS IgG. The clinical sites used a combination of unbiased enrollment and pre-screening for anti-spike and/or anti-nucleocapsid antibodies to achieve sufficient numbers of participants within each serostatus. 506 participants who received the second dose of vaccine on day 28-30 for Covaxin^®^ or day 84-91 for Covishield™, completed all visits until 2 weeks post the second dose of vaccine and did not have symptomatic RT-PCR-confirmed SARS-CoV-2 infection atleast until 2 weeks post the second dose of vaccine, formed the per-protocol population. The 4 study groups, classified based on baseline serostatus and vaccine received, were similar for age and gender across the full cohort (p=0.7534 and 0.8931 respectively) as well as within each cohort (Table 1). The numbers of participants with RT-PCT prior COVID infection were small in each cohort, but did not vary significantly between the study groups within each cohort.

**Table 1.**
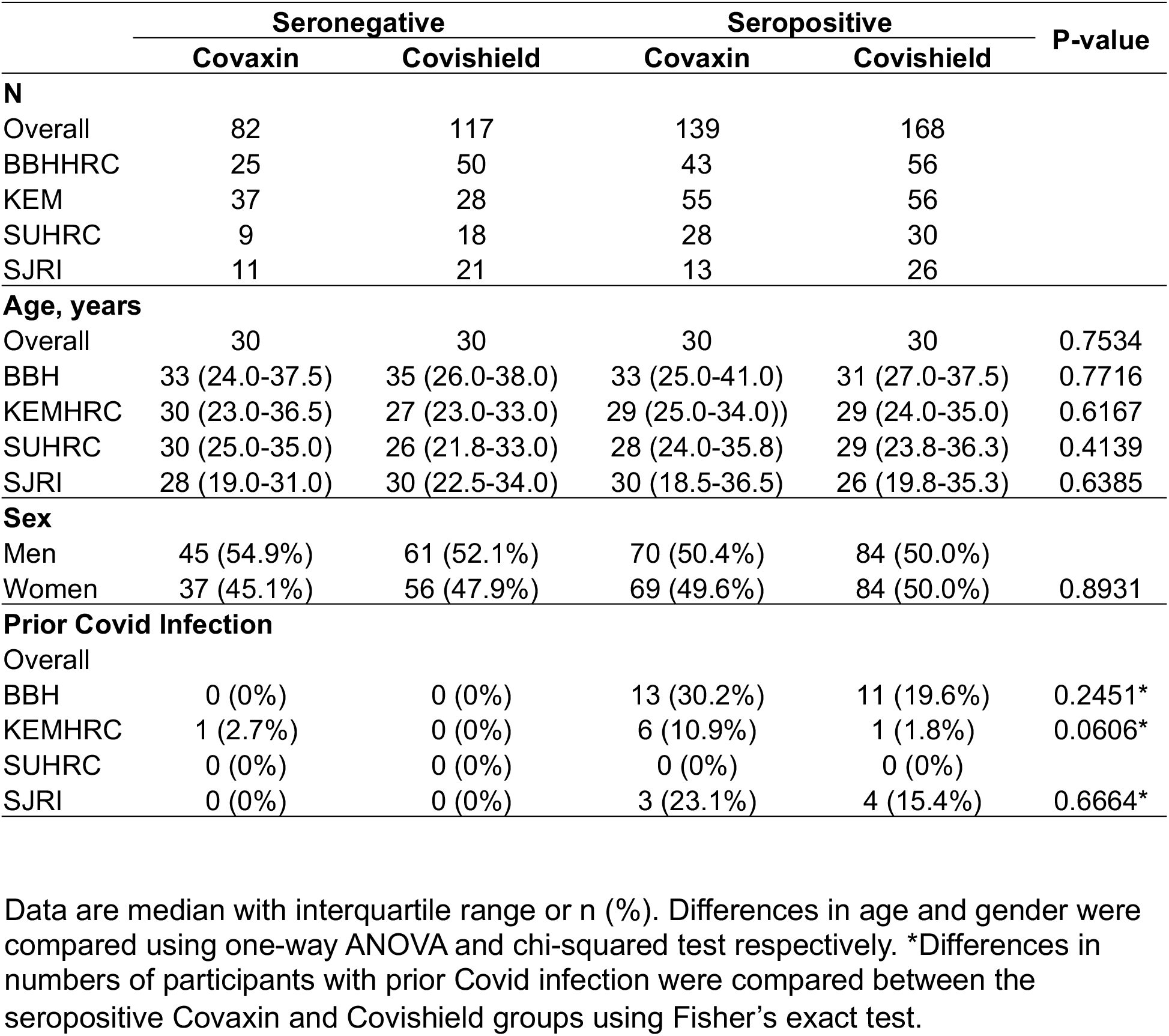
Demographic characteristics of the per-protocol population.

**Figure 1.**
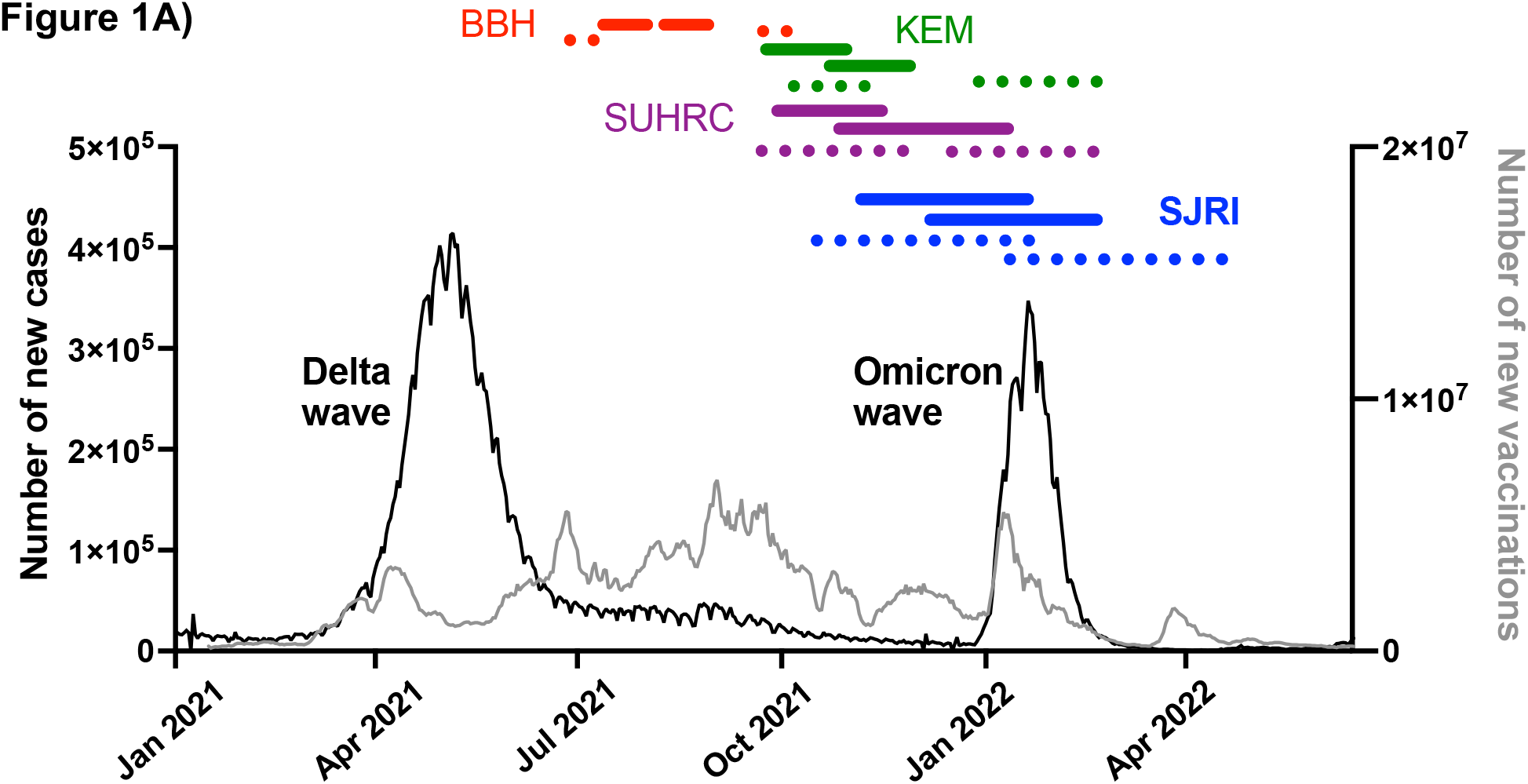
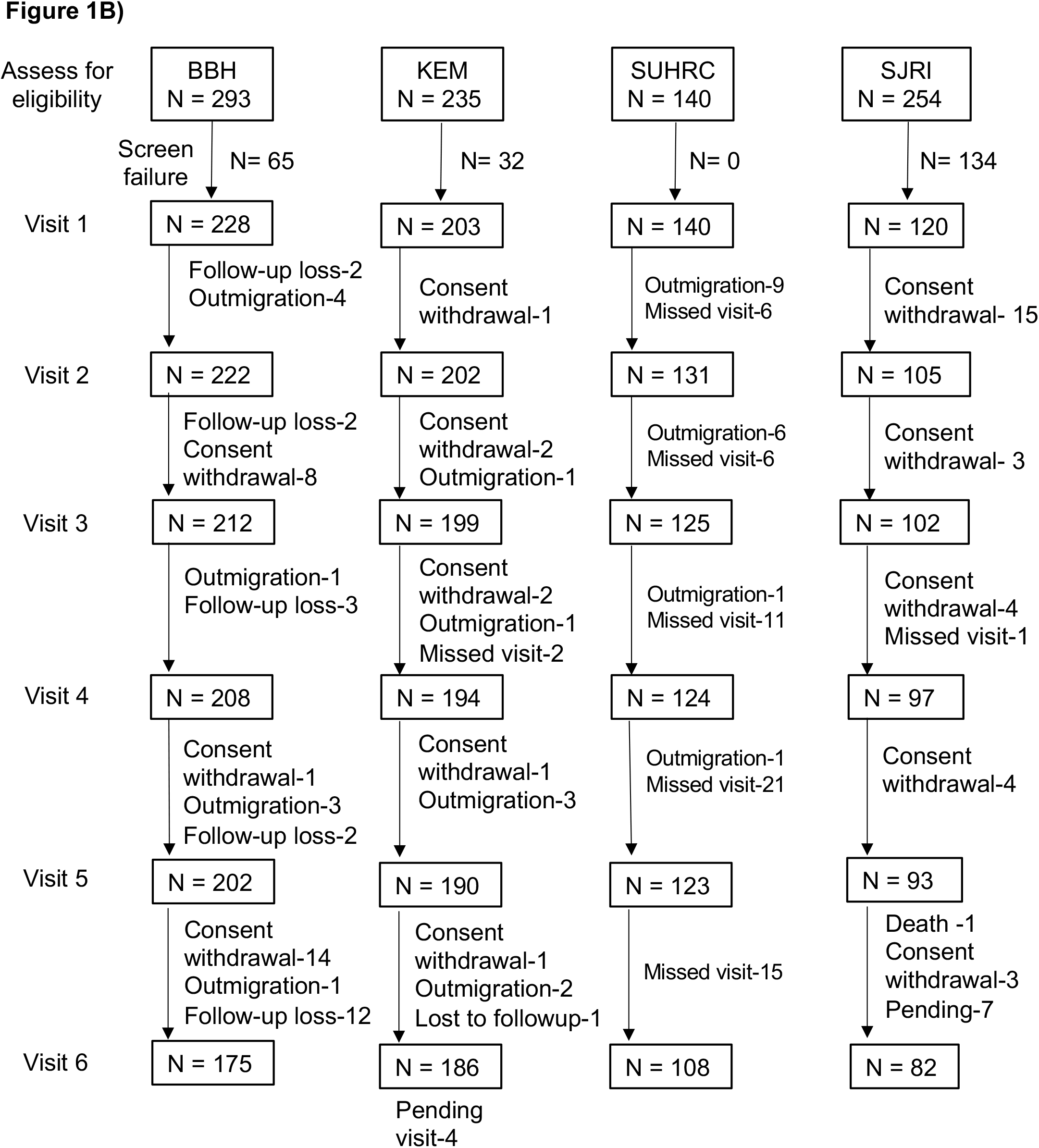
Trial Profile. A) Recruitment schedules across the sites. Horizontal colored lines show the time span during which the first and second doses of vaccine were administered in each site. Solid lines represent the first and second dose of Covaxin and dotted lines represent the first and second dose of Covishield. B) Participant recruitment and follow-up in each of the cohorts.

### Seroconversion rates following vaccination in the per-protocol population

Following vaccination, each dose of vaccine led to a statistically significant increase in the magnitude of anti-spike antibodies over baseline in both seronegative individuals and seropositive individuals (Figure 2A, Supplementary Table 2), as measured using the DiaSorin TrimericS assay platform^12^. At all the timepoints tested, geometric mean titers (GMTs) in the Covishield™ arm were higher than in the Covaxin^®^ arm. We therefore compared seroconversion rates between the Covaxin^®^ and Covishield™ arms, within each baseline serostatus group. Seroconversion was defined as change in titer from <33.8 BAU/ml (manufacturer-specified assay cut off for seropositivity) to >33.8 BAU/ml or an increase in titer by atleast 2-fold if pre-vaccine titer was already >33.8 BAU/ml.

**Figure 2.**
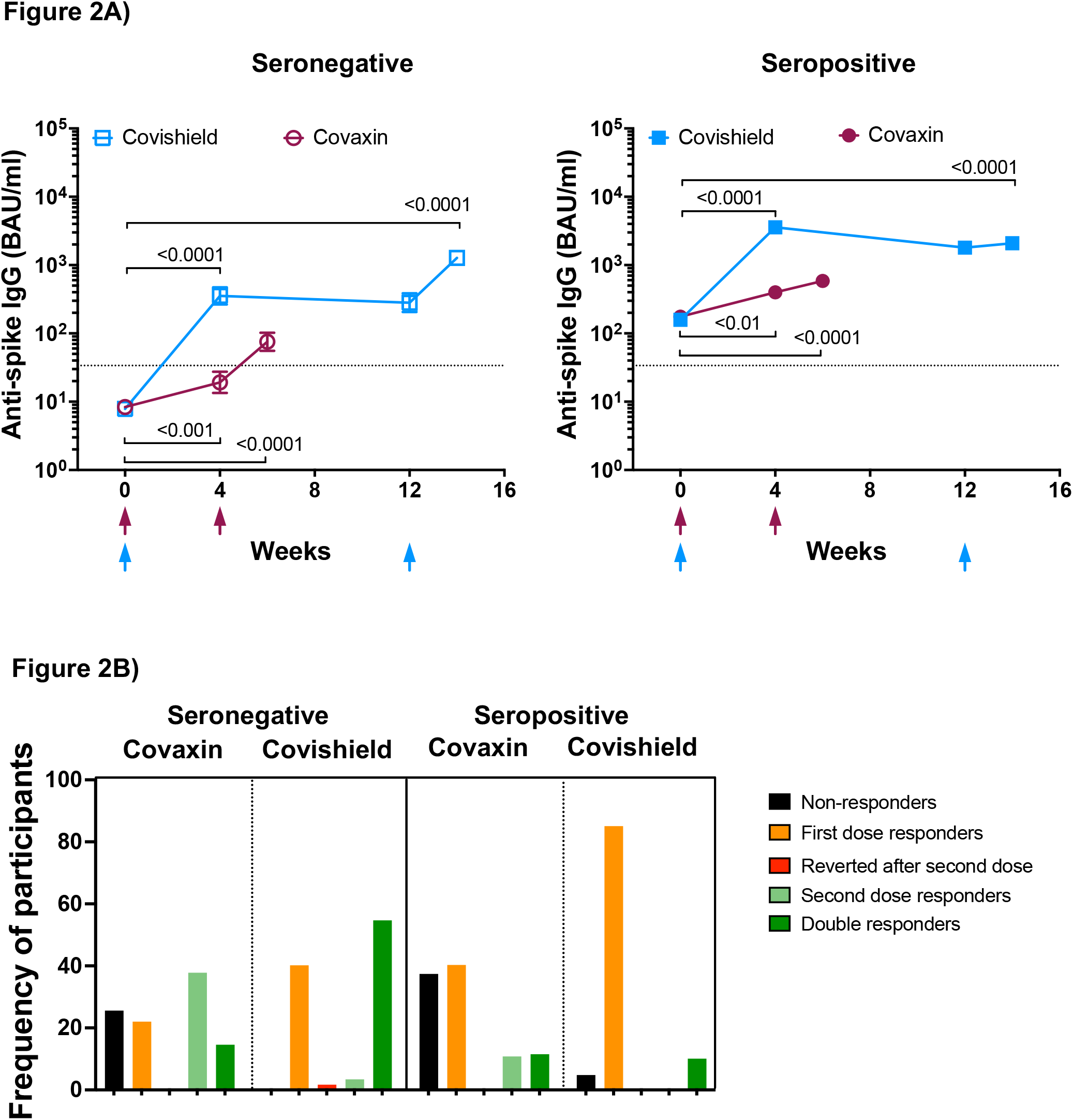
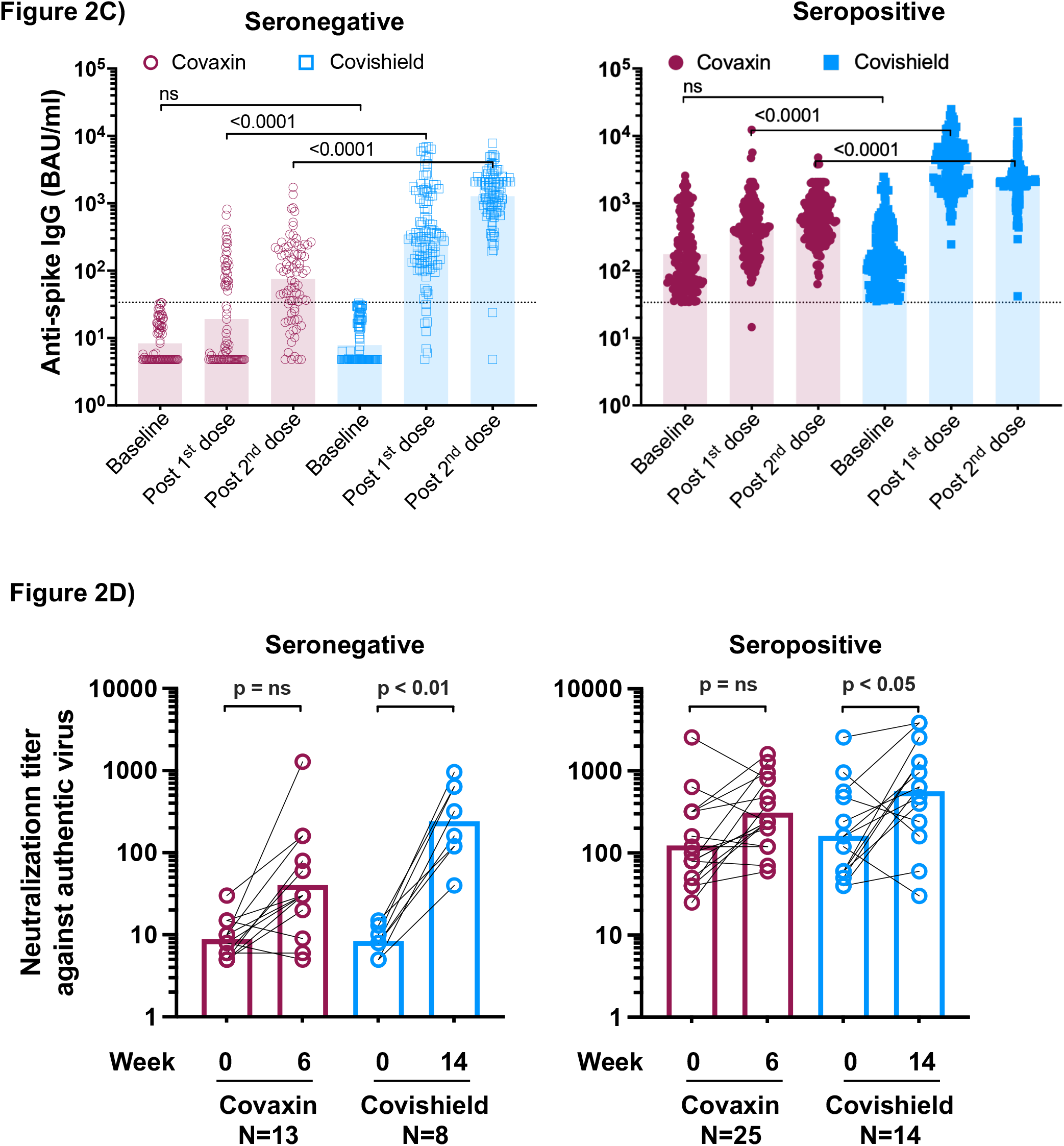
Antibody responses following vaccination in the per-protocol population from all cohorts. Antibody responses against ancestral spike were measured on the Diasorin TrimericS platform in the per-protocol population across the full cohort. Participants were classified as either seronegative (n=82 for Covaxin, open red circles; 117 for Covishield, open blue squares) or seropositive (n = 139 for Covaxin, filled red circles; 168 for Covishield, filled blue squares) at baseline using the assay cut-off of 33.8 BAU/ml (horizontal dotted line). A) Data are shown as geometric mean ± 95% CI. Within-group comparisons to the baseline were assessed using a two-way ANOVA with Tukey’s correction for multiple comparisons. B) Bar graphs show seroconversion rates. Seroconversion was defined as change from <33.8 to >33.8 for those with titers <33.8 BAU/ml prior to the vaccine dose. For those with titers >33.8 BAU/ml, seroconversion was defined as increase in titer by atleast 2-fold following the vaccine administration. C) Between group comparisons after each vaccine dose were made using two-ANOVA with Sidak’s correction. D) Neutralizing responses were measured as inhibition of cytopathic effects by Italian strain INMI1 on VeroE6 cells in a microneutralization assay (see Supp Fig 3A). Participants were classified as either seronegative or positive using a cut-off of 33.8 BAU/ml for ancestral spike binding on the TrimericS platform. Samples from baseline and 2-weeks post the second dose of vaccine were tested. Their corresponding antibody trajectories are shown in Supp figs. 3C and D. Statistical comparisons were made using two-way ANOVA with Sidak’s correction. For all statistical analyses, p-values >0.05 are shown as ns (not significant).

In the baseline seronegative population, seroconversion rates in the Covaxin^®^ arm were 36.6% and 74.4% following the first and second doses of vaccine, respectively (p<0.0001) (Figure 2B, Table 2 and Supplementary Figure 1). Of 36.6% participants who responded to the first dose of Covaxin^®^, only 14.6% also responded to the second dose. 37.8% participants did not show significant responses after the first dose and seroconverted only after the second dose. 25.6% participants classified as non-responders appeared to show detectable antibody responses after the second dose, but did not cross the seropositivity cut-off of the assay. In the seronegative Covishield™ arm, seropositivity rates were 93.2%, and 98.3% following the first and second doses respectively (p<0.0001). The differences in seroconversion rates between the two vaccines in seronegative individuals were significant in the overall cohort both after the first and second dose (p<0.0001) and in 2 of the 4 sites after the second dose (Table 2; see also Figure 4).

**Table 2.**
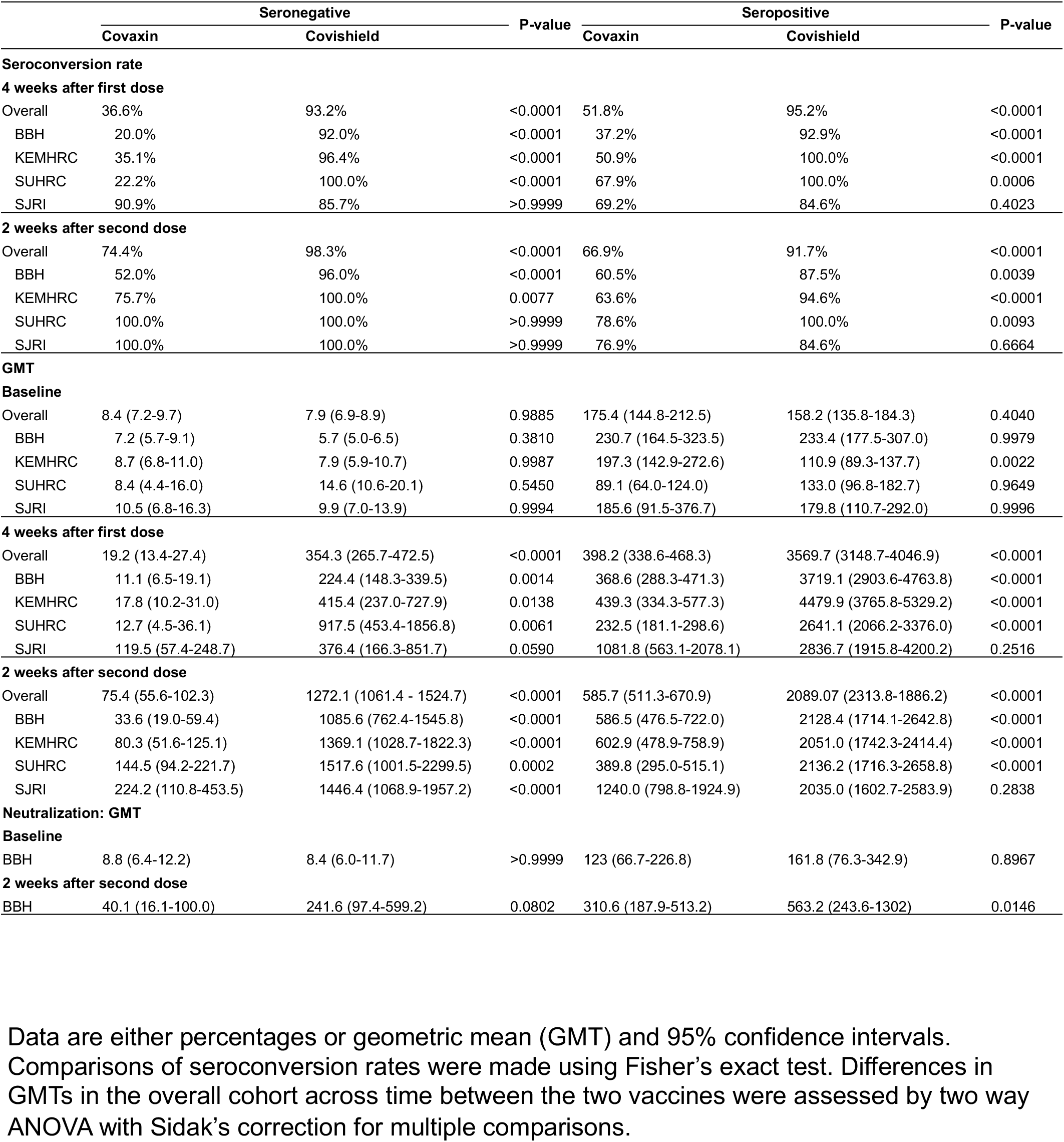
Vaccine-elicited antibody responses in the per-protocol population.

In the baseline seropositive group, Covaxin^®^ administration led to an increase in antibody titer by at least 2-fold over baseline in 51.8% and 66.9% participants, following the first dose and second dose respectively (p<0.0001, Table 2). Among the 51.8% first-dose responders, only 11.5% participants also additionally responded to the second dose of vaccine while the other 40.3% participants did not show a further increase in titers after the second dose of vaccine. 10.8% participants responded only to the second dose of vaccine and 37.4% participants did not respond to either dose (Figure 2B and supplementary figure 1A). In the Covishield™ group, antibody titers increased atleast 2-fold over baseline in 95.2%, and 91.7% participants after the first and second doses respectively (p<0.0001, Table 2). 10.1% participants responded to both vaccine doses, 85.1% participants responded only to the first dose and 4.8% did not respond to either dose. The differences in seroconversion rates between the two vaccines after the first and second doses were significant in the overall cohort (p<0.0001). In both vaccine arms, non-responsiveness to vaccination was associated with higher pre-vaccination antibody titers (Supplementary Figure 1B).

Though trends in vaccine- and serostatus-wise immunogenicity outcomes remained the same within each of the four cohorts, the magnitude of differences between study arms were impacted by the recruitment period of the cohorts (Figure 1A). Notably, the two cohorts with significant differences in seroconversion rates between the vaccines had sampled participants in the Covaxin^®^ arm prior to the Omicron wave, while the other two cohorts with higher seroconversion rates for Covaxin^®^ had dosed and/or sampled the Covaxin^®^ arm during the Omicron wave. The trends in differences in cohort-wise immunogenicity between the two vaccines in seropositive individuals was similar to that of the seronegative individuals. Cohorts that were vaccinated and sampled soon after the delta wave had lower seroconversion rates for Covaxin^®^ than for Covishield™. Cohorts that were vaccinated and sampled overlapping the Omicron wave displayed comparable immunogenicity between the two vaccines (Table 2; Supplementary Figure 2).

### Magnitude of anti-spike antibodies following vaccination in the per-protocol population

The geometric mean of the antibody titer (GMT) in the seronegative Covaxin^®^ group increased from 8.4 BAU/ml at baseline to 19.2 BAU/ml and 75.4 BAU/ml following the first dose and second doses, respectively (Figure 2C and Table 2). In the seronegative Covishield™ group, titers increased from 7.9 BAU/ml to 354.3 BAU/ml and 1272.1 BAU/ml following the first and second doses, respectively (p<0.0001 after both doses). The GMTs in the Covishield™ arm were significantly higher than in the Covaxin^®^ arm at both time points (p<0.0001).

The GMT in the seropositive Covaxin^®^ group increased from 175.4 BAU/ml to 398.2 BAU/ml and 585.7 BAU/ml following the first dose and second doses of respectively (p=0.0037 and <0.0001, Figure 2C). In the seropositive Covishield™ arm, GMT increased from 158.21 BAU/ml to 3569.69 BAU/ml and 2089.07 BAU/ml (p<0.0001). Despite waning of antibody titers between weeks 4 and 12, a second dose of Covishield™ at week 12 did not restore the antibody levels seen after the first dose of vaccine. The difference in titers between the vaccines was significant after each dose (p<0.0001).

Considering the pre-vaccination titers in the seropositive group as a reference for convalescent titers, GMT after first and second dose of Covishield™ in seronegative individuals were 2.1 and 7.6 -fold higher, whereas two doses of Covaxin^®^ in seronegative individuals remained 2.2-fold lower than that of the convalescents.

### Magnitude of anti-spike antibodies within each cohort

In the seronegative Covaxin^®^ arm, there was a trend towards increasing post-vaccine antibody titers in the cohorts that started recruitment later and overlapped with the Omicron wave. However, the inter-cohort differences in the GMTs were not statistically significant, likely due to insufficient sample size in some of the cohorts. (Supplementary Figure 2).

In the seronegative Covishield™ arm, there were minor but statistically significant differences in baseline titers, suggesting that some of the participants who were classified as seronegative by the TrimericS platform may have been previously exposed and antibody levels may have waned off at the time of recruitment. Nevertheless, all cohorts reached similar titers after the first dose. Significant differences again arose at week 12 due to differential antibody kinetics across cohorts. At week 12, the BBH cohort that was sampled prior to the Omicron wave had the lowest titer at 90.5 BAU/ml, KEMHRC and SUHRC cohorts that received the second dose and were sampled during the Omicron wave were intermediate at 366.5 and 611.2 BAU/ml and the SJRI cohort that was positively impacted by the Omicron wave during and between the first and second doses had the highest titer at 1522.1 BAU/ml. The SJRI cohort with first dose vaccination during the Omicron wave showed increase in antibody titers between weeks 4 and 12, whereas the other three cohorts showed stable or declining titers. These differences in week 12 were nullified at week 14 due to the second dose of vaccine, with all cohorts reaching similar antibody titers irrespective of their exposure to infection and/or vaccination.

In the seropositive Covaxin^®^ group, between-cohort differences in titers were in the 2.4-2.6 fold range, some of which were significant. In the seropositive Covishield™ group, the baseline titers showed statistically significant variation across cohorts but normalized after the first dose of vaccination. The differences among the cohorts in exposure to the Omicron wave was not manifested in the antibody responses in the sSeropositive Covishield™ group.

We also measured neutralizing antibodies using the authentic virus at baseline and at two weeks post the second dose of vaccine. The neutralization titers measured in this assay correlated significantly with the anti-spike IgG and with ACE2 inhibition measured using a MSD based assay (Supplementary Figure 3A). We compared neutralizing antibody titers in a subset of participants from one cohort (BBH) and matched for baseline anti-spike binding antibody titers between the two vaccine arms and representative of the spike-binding antibody responses in each vaccine arm (Supplementary Figure 3B and C). Covishield™ elicited significant levels of neutralizing antibodies in both seronegative individuals (GMT 8.4 to 241.6; p=0.0085) and seropositive individuals (GMT 161.8 to 563.2; p=0.0263). However, Covaxin^®^ failed to elicit statistically significant increases in GMT in the Covaxin^®^ group, although the GMT increased from 8.8 to 40.1 in seronegative individuals and 123.0 to 310.6 in seropositive individuals (Figure 2D).

### T cell responses following vaccination

Across all vaccine platforms against SARS-CoV-2, spike-specific antibodies wane considerably over a six-month period and it has been proposed that adaptive immune memory, especially T cells, might be sufficient to ward off symptomatic infection and severe disease. We therefore measured the frequency of spike-specific CD4 and CD8 T cell responses at baseline and at 2 weeks post the second vaccine dose in a subset of participants from the earliest recruited cohort (BBH). Supplementary Figure 4 shows representative gating strategy for viable responding CD4 and CD8 cells and intracellular cytokine staining in the presence and absence of specific peptides and antibody trajectories for samples included in the ICS assay. In concordance with the spike antibody-based classification of participants into baseline seronegative individuals and seropositive individuals, there were significantly higher frequency of CD4 T cells against an ancestral spike peptide pool in the baseline seropositive group than in the seronegative group (Supplementary Figure 5). However, in the baseline samples there were no significant differences between seronegative individuals and seropositive individuals in total CD4 or CD8 T cell responses against a pool of nucleocapsid and matrix peptides or in the frequencies of CD8 T cells against spike. The frequency of spike-specific and nucleocapsid-specific IL2-producing CD4 T cells were significantly associated with the magnitude of spike-binding antibodies at baseline.

In seronegative individuals, both Covaxin^®^ and Covishield™ led to increases in median spike-specific CD4 T cells following vaccination (0.002 to 0.01% and 0.005 to 0.07% respectively, Figure 3A), though the fold change was statistically significant only in the Covishield™ arm (p=0.0022). Given the lower seroconversion rate in the seronegative Covaxin^®^ arm, we split this group further as antibody-responders vs non-responders (Supplementary Figure 6). As expected, significant spike-specific CD4 T cells were observed in the Covaxin^®^ antibody responder group (0.002% to 0.024%, p< 0.05). Spike-specific CD8 T cell responses were only detected in the Covishield™ arm (0.002% to 0.035%, p<0.05) but not in the Covaxin^®^ arm. When analyzed for individual cytokines, Covishield™ elicited significant IFNγ, IL2 and TNF responses in CD4 T cells, whereas Covaxin^®^ elicited IL2 and TNF responses, however these did not reach statistical significance (Supplementary Figure 6C). Despite elicitation of nucleocapsid antibodies by Covaxin^®^, we did not detect a significant expansion of the CD4 T cells against the N+M peptide pool.

**Figure 3.**
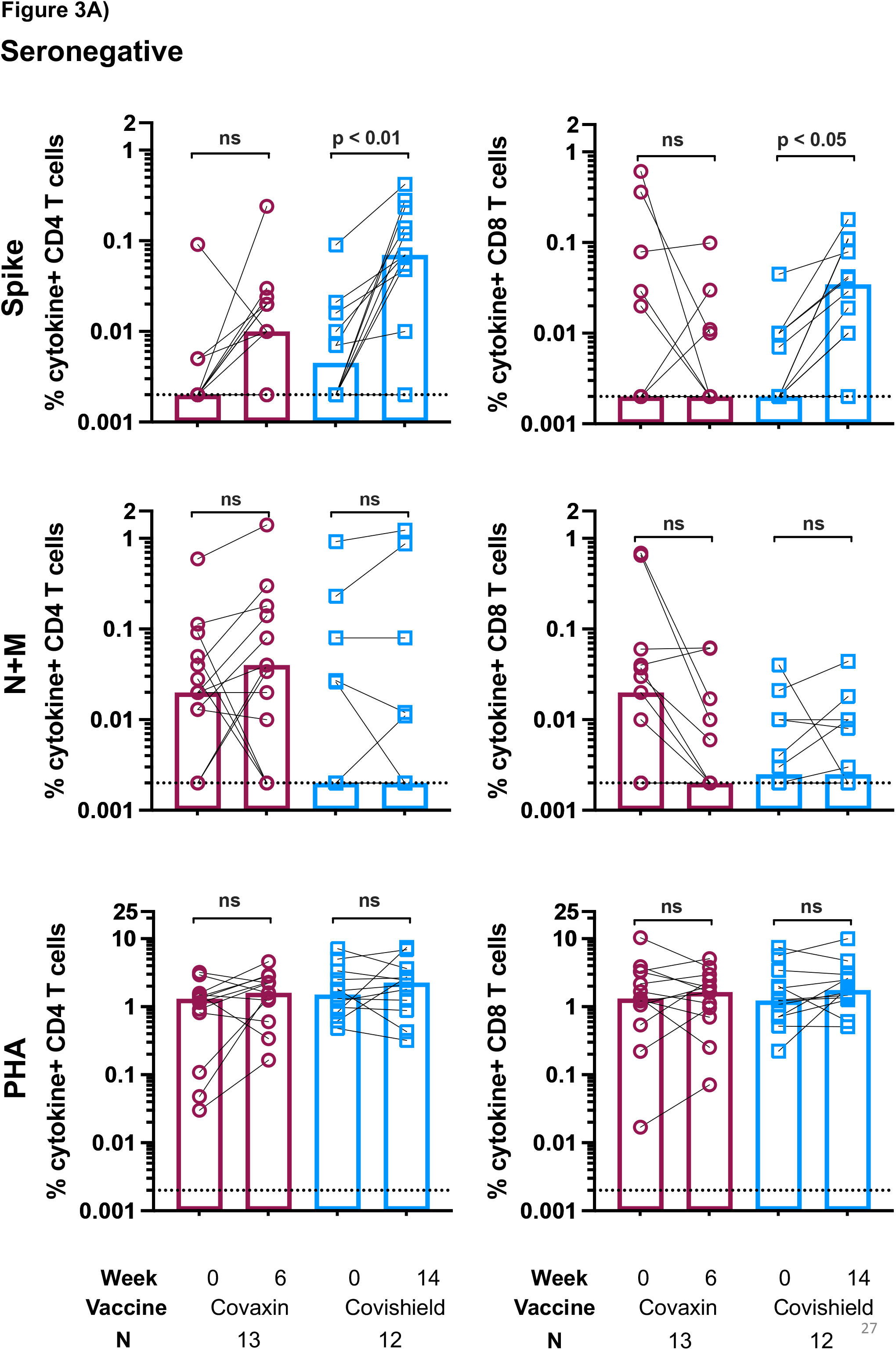
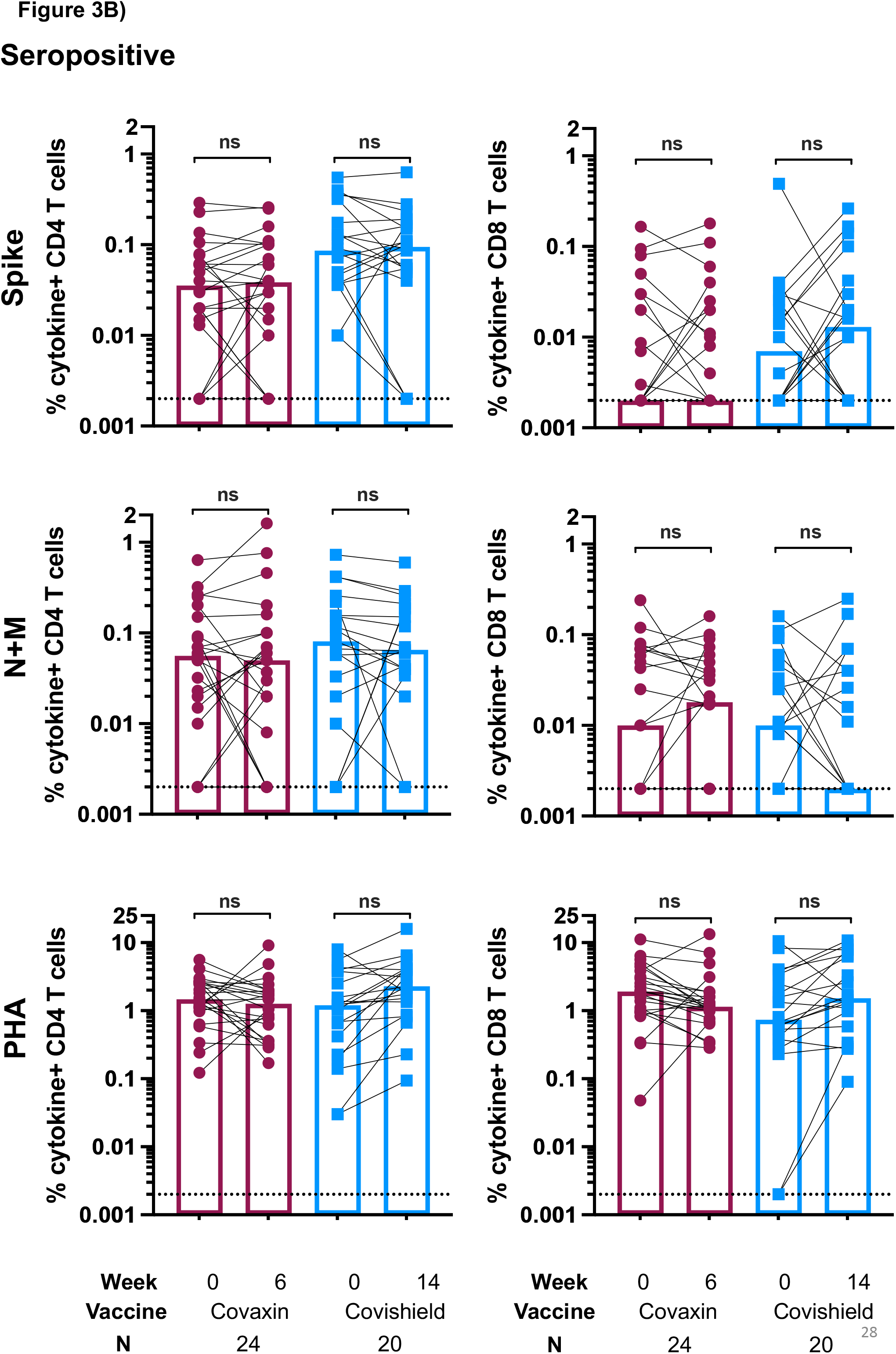
Vaccine-induced T cell responses. Frequency of spike-specific CD4 or CD8 T cells expressing at least one cytokine amongst IFNγ, IL2 and TNF in response to stimulation with either spike peptide pool, mixture of nucleocapsid and matrix peptide pools or PHA, following vaccination in A) Seronegative Covaxin (red) or Covishield (blue) B) Seropositive Covaxin (red) or Covishield (blue). Frequencies are shown after background subtraction from the unstimulated control. Differences between pre- and post-vaccination responses were compared using Kruskal-Wallis test with Dunn’s correction. p-values >0.05 are shown as ns (not significant).

In seropositive individuals, we did not detect a significant increase in the frequency of spike-specific CD4 or CD8 T cells with either vaccine (Figure 3B). In both vaccine arms, there was a trend for group medians of cytokine-producing T cells in antibody non-responders to decrease following vaccination and in responders to either marginally increase or remain stable (Supplementary Figures 7A and B). When the cytokines were analyzed individually, there appeared to be trend for spike-specific CD8 T cell responses to increase in the seropositive Covishield™ arm (Supplementary Figure 7C).

### Impact of the Omicron wave on binding antibody responses

Due to exposure to the Omicron wave, durability of vaccine-elicited responses could not be assessed. In the earliest cohort (BBH), however, data from the predominantly asymptomatic and symptomatic infection-free data was available from baseline until 6 months post first-dose of vaccine. Despite low peak antibody responses at week 6, the seronegative Covaxin^®^ group showed fairly stable antibody responses until week 12 and week 25, hovering around the seropositivity cut-off at 33.8 BAU/ml (Table 3 and Figure 4A). Similar trends were observed when samples were tested on the MSD platform with a higher dynamic range. Notably, week 25 was sampled during the Omicron wave in January 2022 and participants may have been variably sampled either prior to or after asymptomatic Omicron infection. In support of this, the individual spike and nucleocapsid antibody trajectories between weeks 12 and 40 suggest that almost all participants in the seronegative Covaxin^®^ group may have been asymptomatically infected during this period. Though Covaxin^®^ administration led to increase in both spike and nucleocapsid antibodies at weeks 4 and 6, the increase in titer at week 40 was much higher. This suggests that both spike and nucleocapsid antibodies were sub-optimally induced by the Covaxin^®^ formulation. The seronegative Covishield™ group was predominantly sampled during December 2021, several days before the Omicron wave was recorded in India, and therefore it is not clear if the 4.1-fold decline in antibody titers between the peak antibody response at week 14 and week 25 is truly reflective of antibody waning. Similar to the Covaxin^®^ arm, titers increased 3.8-fold between week 25 and week 40 in the seronegative Covishield™ arm.

**Figure 4.**
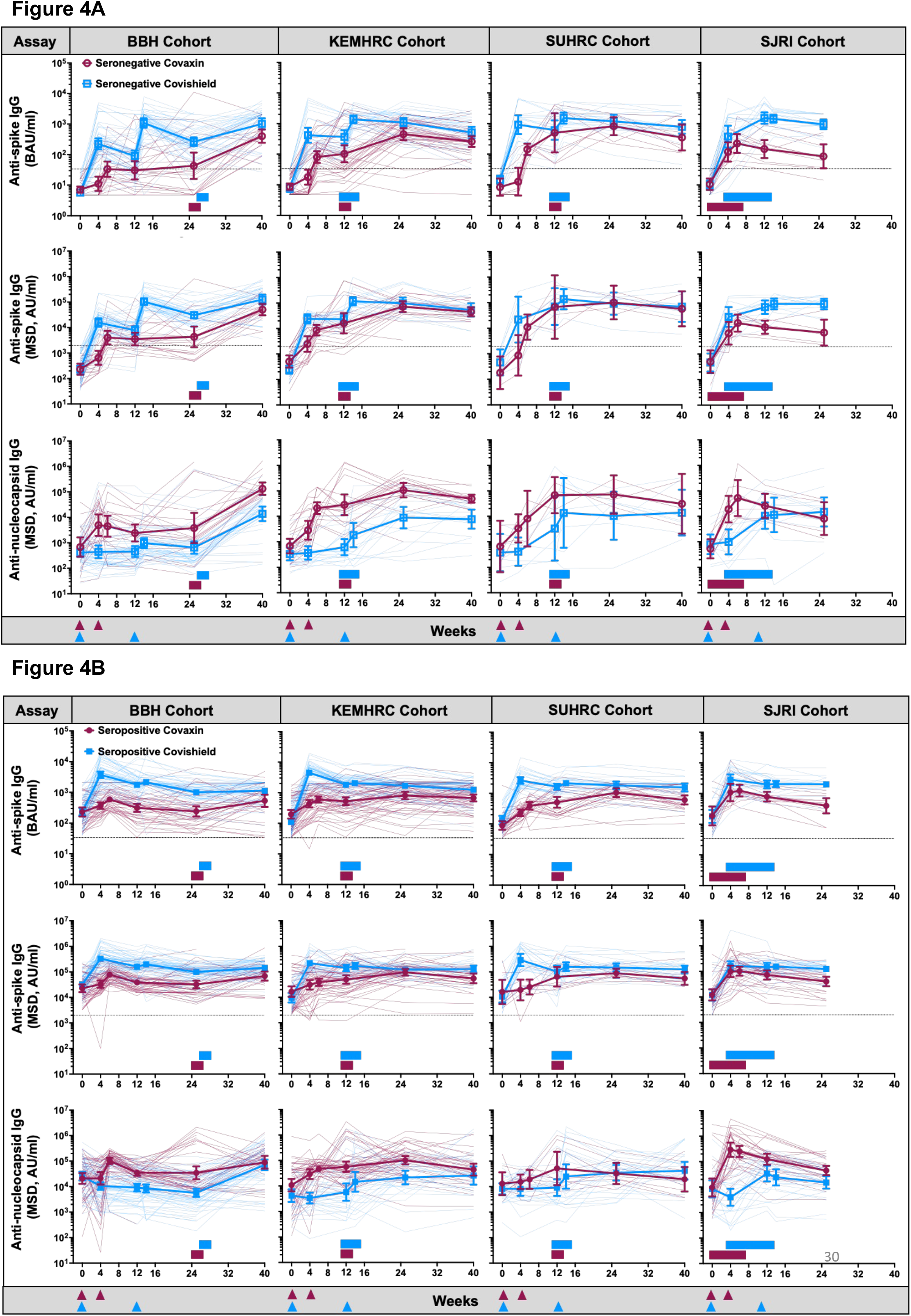
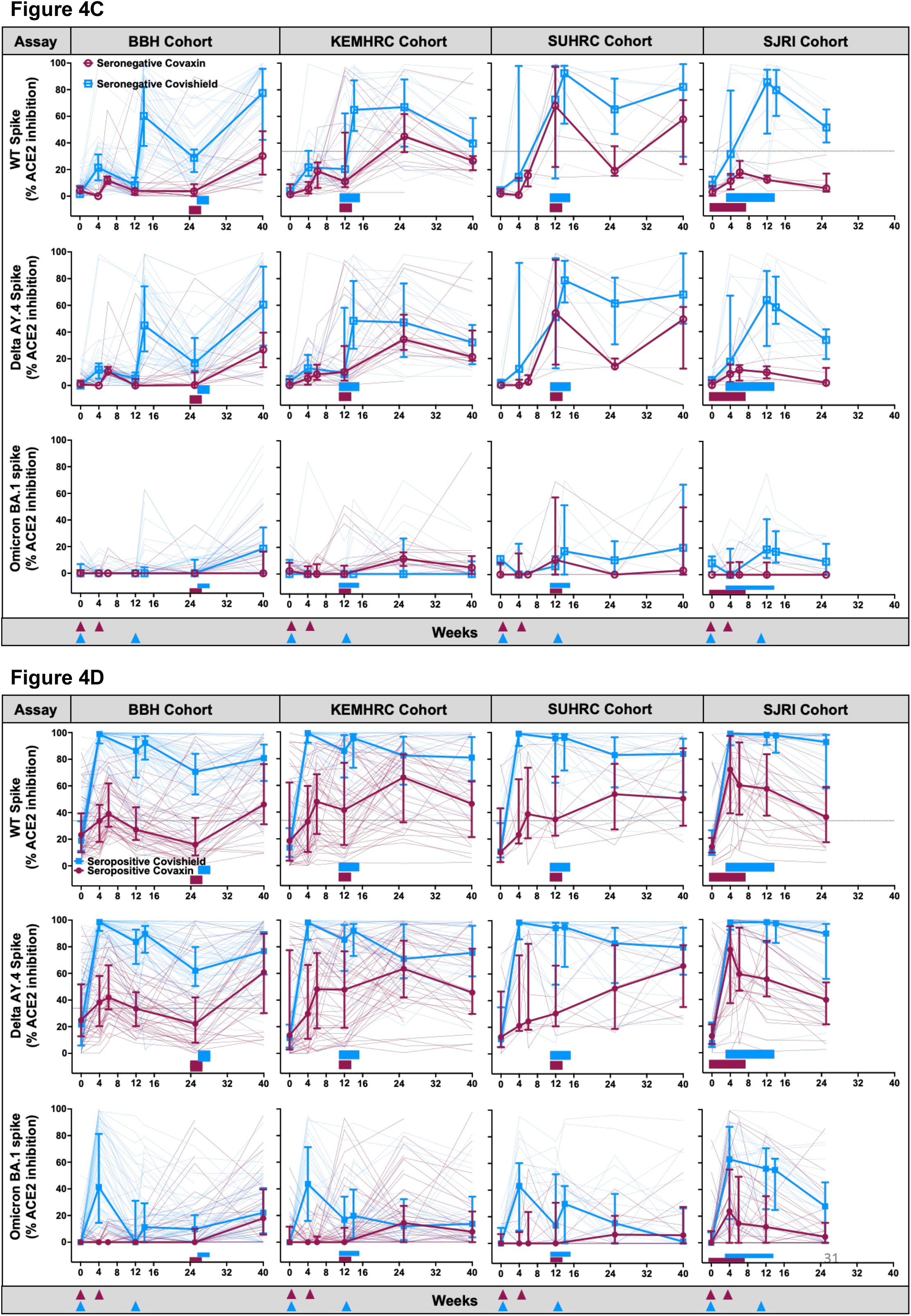
Durability and breadth of antibody responses following vaccination and exposure to the Omicron wave. A, B) Durability of antibody responses against spike protein (both Diasorin TrimericS and MSD platforms) and nucleocapsid protein (MSD) are shown for each cohort as geometric mean ± 95% CI for seronegative individuals (A) and seropositive individuals (B). C, D) Inhibition of ACE2 binding to the indicated spike proteins is shown as medians with interquartile range for seronegative individuals (C) and seropositive individuals (D). Trajectories for individual participants are shown as thin background lines. Horizontal bars (red - Covaxin^®^, blue - Covishield) within each graph show timepoints in which sampling was performed during the Omicron wave

**Table 3.**
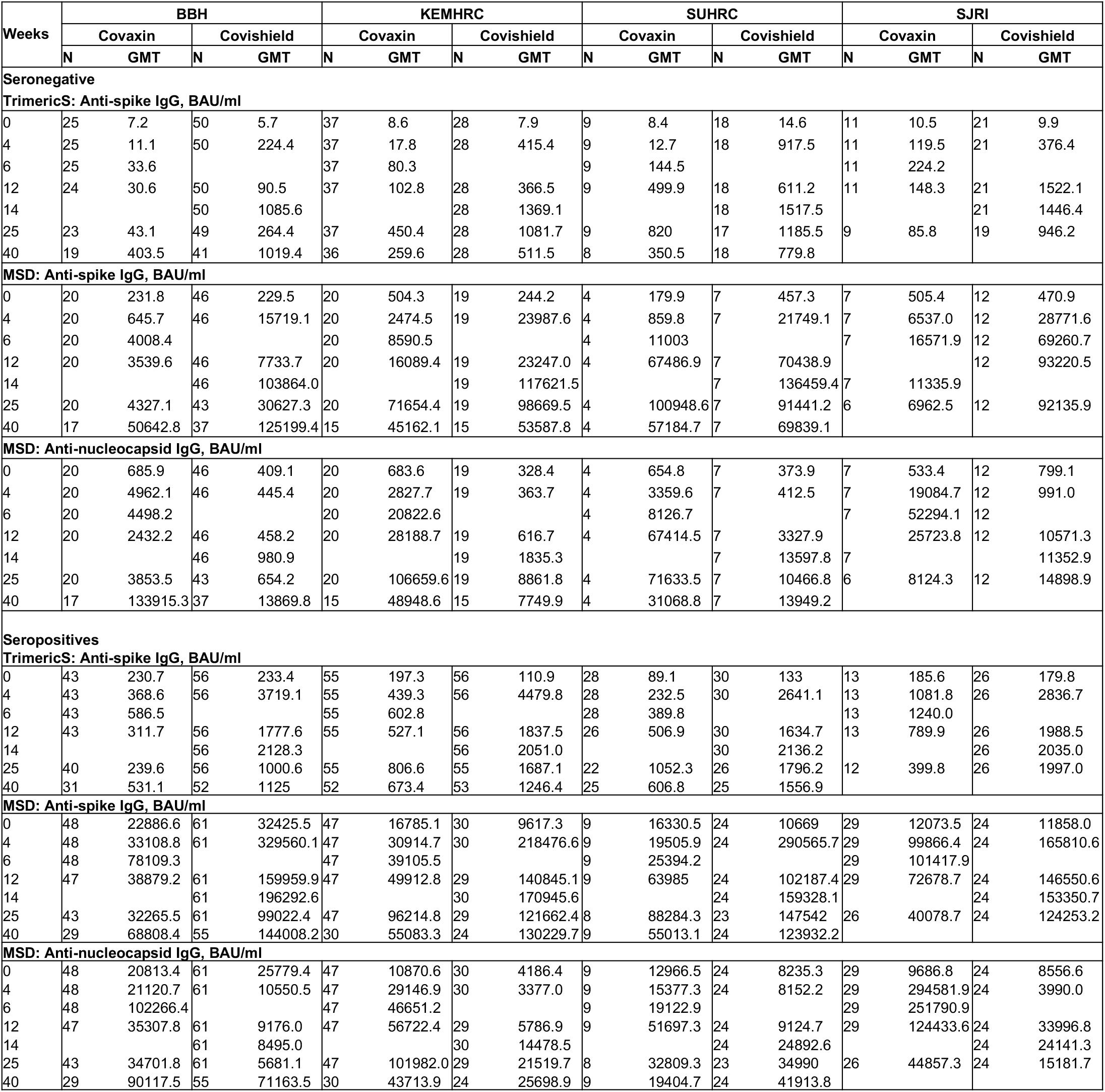
Antibody titers in all cohorts across all study timepoints. Data are shown as N and geometric mean titers

| Weeks | BBH |  |  |  | KEMHRC |  |  |  | SUHRC |  |  |  | SJRI |  |  |  |
| --- | --- | --- | --- | --- | --- | --- | --- | --- | --- | --- | --- | --- | --- | --- | --- | --- |
|  | Covaxin |  | Covishield |  | Covaxin |  | Covishield |  | Covaxin |  | Covishield |  | Covaxin |  | Covishield |  |
|  | N | GMT | N | GMT | N | GMT | N | GMT | N | GMT | N | GMT | N | GMT | N | GMT |
| Seronegative |  |  |  |  |  |  |  |  |  |  |  |  |  |  |  |  |
| TrimericS: Anti-spike IgG, BAU/ml |  |  |  |  |  |  |  |  |  |  |  |  |  |  |  |  |
| 0 | 25 | 7.2 | 50 | 5.7 | 37 | 8.6 | 28 | 7.9 | 9 | 8.4 | 18 | 14.6 | 11 | 10.5 | 21 | 9.9 |
| 4 | 25 | 11.1 | 50 | 224.4 | 37 | 17.8 | 28 | 415.4 | 9 | 12.7 | 18 | 917.5 | 11 | 119.5 | 21 | 376.4 |
| 6 | 25 | 33.6 |  |  | 37 | 80.3 |  |  | 9 | 144.5 |  |  | 11 | 224.2 |  |  |
| 12 | 24 | 30.6 | 50 | 90.5 | 37 | 102.8 | 28 | 366.5 | 9 | 499.9 | 18 | 611.2 | 11 | 148.3 | 21 | 1522.1 |
| 14 |  |  | 50 | 1085.6 |  |  | 28 | 1369.1 |  |  | 18 | 1517.5 |  |  | 21 | 1446.4 |
| 25 | 23 | 43.1 | 49 | 264.4 | 37 | 450.4 | 28 | 1081.7 | 9 | 820 | 17 | 1185.5 | 9 | 85.8 | 19 | 946.2 |
| 40 | 19 | 403.5 | 41 | 1019.4 | 36 | 259.6 | 28 | 511.5 | 8 | 350.5 | 18 | 779.8 |  |  |  |  |
| MSD: Anti-spike IgG, BAU/ml |  |  |  |  |  |  |  |  |  |  |  |  |  |  |  |  |
| 0 | 20 | 231.8 | 46 | 229.5 | 20 | 504.3 | 19 | 244.2 | 4 | 179.9 | 7 | 457.3 | 7 | 505.4 | 12 | 470.9 |
| 4 | 20 | 645.7 | 46 | 15719.1 | 20 | 2474.5 | 19 | 23987.6 | 4 | 859.8 | 7 | 21749.1 | 7 | 6537.0 | 12 | 28771.6 |
| 6 | 20 | 4008.4 |  |  | 20 | 8590.5 |  |  | 4 | 11003 |  |  | 7 | 16571.9 | 12 | 69260.7 |
| 12 | 20 | 3539.6 | 46 | 7733.7 | 20 | 16089.4 | 19 | 23247.0 | 4 | 67486.9 | 7 | 70438.9 |  |  | 12 | 93220.5 |
| 14 |  |  | 46 | 103864.0 |  |  | 19 | 117621.5 |  |  | 7 | 136459.4 | 7 | 11335.9 |  |  |
| 25 | 20 | 4327.1 | 43 | 30627.3 | 20 | 71654.4 | 19 | 98669.5 | 4 | 100948.6 | 7 | 91441.2 | 6 | 6962.5 | 12 | 92135.9 |
| 40 | 17 | 50642.8 | 37 | 125199.4 | 15 | 45162.1 | 15 | 53587.8 | 4 | 57184.7 | 7 | 69839.1 |  |  |  |  |
| MSD: Anti-nucleocapsid IgG, BAU/ml |  |  |  |  |  |  |  |  |  |  |  |  |  |  |  |  |
| 0 | 20 | 685.9 | 46 | 409.1 | 20 | 683.6 | 19 | 328.4 | 4 | 654.8 | 7 | 373.9 | 7 | 533.4 | 12 | 799.1 |
| 4 | 20 | 4962.1 | 46 | 445.4 | 20 | 2827.7 | 19 | 363.7 | 4 | 3359.6 | 7 | 412.5 | 7 | 19084.7 | 12 | 991.0 |
| 6 | 20 | 4498.2 |  |  | 20 | 20822.6 |  |  | 4 | 8126.7 |  |  | 7 | 52294.1 | 12 |  |
| 12 | 20 | 2432.2 | 46 | 458.2 | 20 | 28188.7 | 19 | 616.7 | 4 | 67414.5 | 7 | 3327.9 |  | 25723.8 | 12 | 10571.3 |
| 14 |  |  | 46 | 980.9 |  |  | 19 | 1835.3 |  |  | 7 | 13597.8 | 7 |  |  | 11352.9 |
| 25 | 20 | 3853.5 | 43 | 654.2 | 20 | 106659.6 | 19 | 8861.8 | 4 | 71633.5 | 7 | 10466.8 | 6 | 8124.3 | 12 | 14898.9 |
| 40 | 17 | 133915.3 | 37 | 13869.8 | 15 | 48948.6 | 15 | 7749.9 | 4 | 31068.8 | 7 | 13949.2 |  |  |  |  |
| Seropositives |  |  |  |  |  |  |  |  |  |  |  |  |  |  |  |  |
| TrimericS: Anti-spike IgG, BAU/ml |  |  |  |  |  |  |  |  |  |  |  |  |  |  |  |  |
| 0 | 43 | 230.7 | 56 | 233.4 | 55 | 197.3 | 56 | 110.9 | 28 | 89.1 | 30 | 133 | 13 | 185.6 | 26 | 179.8 |
| 4 | 43 | 368.6 | 56 | 3719.1 | 55 | 439.3 | 56 | 4479.8 | 28 | 232.5 | 30 | 2641.1 | 13 | 1081.8 | 26 | 2836.7 |
| 6 | 43 | 586.5 |  |  | 55 | 602.8 |  |  | 28 | 389.8 |  |  | 13 | 1240.0 |  |  |
| 12 | 43 | 311.7 | 56 | 1777.6 | 55 | 527.1 | 56 | 1837.5 | 26 | 506.9 | 30 | 1634.7 | 13 | 789.9 | 26 | 1988.5 |
| 14 |  |  | 56 | 2128.3 |  |  | 56 | 2051.0 |  |  | 30 | 2136.2 |  |  | 26 | 2035.0 |
| 25 | 40 | 239.6 | 56 | 1000.6 | 55 | 806.6 | 55 | 1687.1 | 22 | 1052.3 | 26 | 1796.2 | 12 | 399.8 | 26 | 1997.0 |
| 40 | 31 | 531.1 | 52 | 1125 | 52 | 673.4 | 53 | 1246.4 | 25 | 606.8 | 25 | 1556.9 |  |  |  |  |
| MSD: Anti-spike IgG, BAU/ml |  |  |  |  |  |  |  |  |  |  |  |  |  |  |  |  |
| 0 | 48 | 22886.6 | 61 | 32425.5 | 47 | 16785.1 | 30 | 9617.3 | 9 | 16330.5 | 24 | 10669 | 29 | 12073.5 | 24 | 11858.0 |
| 4 | 48 | 33108.8 | 61 | 329560.1 | 47 | 30914.7 | 30 | 218476.6 | 9 | 19505.9 | 24 | 290565.7 | 29 | 99866.4 | 24 | 165810.6 |
| 6 | 48 | 78109.3 |  |  | 47 | 39105.5 |  |  | 9 | 25394.2 |  |  | 29 | 101417.9 |  |  |
| 12 | 47 | 38879.2 | 61 | 159959.9 | 47 | 49912.8 | 29 | 140845.1 | 9 | 63985 | 24 | 102187.4 | 29 | 72678.7 | 24 | 146550.6 |
| 14 |  |  | 61 | 196292.6 |  |  | 30 | 170945.6 |  |  | 24 | 159328.1 |  |  | 24 | 153350.7 |
| 25 | 43 | 32265.5 | 61 | 99022.4 | 47 | 96214.8 | 29 | 121662.4 | 8 | 88284.3 | 23 | 147542 | 26 | 40078.7 | 24 | 124253.2 |
| 40 | 29 | 68808.4 | 55 | 144008.2 | 30 | 55083.3 | 24 | 130229.7 | 9 | 55013.1 | 24 | 123932.2 |  |  |  |  |
| MSD: Anti-nucleocapsid IgG, BAU/ml |  |  |  |  |  |  |  |  |  |  |  |  |  |  |  |  |
| 0 | 48 | 20813.4 | 61 | 25779.4 | 47 | 10870.6 | 30 | 4186.4 | 9 | 12966.5 | 24 | 8235.3 | 29 | 9686.8 | 24 | 8556.6 |
| 4 | 48 | 21120.7 | 61 | 10550.5 | 47 | 29146.9 | 30 | 3377.0 | 9 | 15377.3 | 24 | 8152.2 | 29 | 294581.9 | 24 | 3990.0 |
| 6 | 48 | 102266.4 |  |  | 47 | 46651.2 |  |  | 9 | 19122.9 |  |  | 29 | 251790.9 |  |  |
| 12 | 47 | 35307.8 | 61 | 9176.0 | 47 | 56722.4 | 29 | 5786.9 | 9 | 51697.3 | 24 | 9124.7 | 29 | 124433.6 | 24 | 33996.8 |
| 14 |  |  | 61 | 8495.0 |  |  | 30 | 14478.5 |  |  | 24 | 24892.6 |  |  | 24 | 24141.3 |
| 25 | 43 | 34701.8 | 61 | 5681.1 | 47 | 101982.0 | 29 | 21519.7 | 8 | 32809.3 | 23 | 34990 | 26 | 44857.3 | 24 | 15181.7 |
| 40 | 29 | 90117.5 | 55 | 71163.5 | 30 | 43713.9 | 24 | 25698.9 | 9 | 19404.7 | 24 | 41913.8 |  |  |  |  |

Sampling in the seropositive groups of the BBH cohort followed the same vaccine-wise trend as in the seronegative group (Figure 4B). Between weeks 6, and 25, titers declined by 2.4-fold in the seropositive Covaxin^®^ group (Table 3). In comparison to the 4.1-fold decline in the seronegative Covishield™ arm, the seropositive Covishield™ arm only showed a 2.1-fold decline in antibody levels between weeks 14 and 25. At week 40, most participants in the seropositive Covishield™ arm had increased nucleocapsid antibodies, with a >10-fold increase in GMT, suggesting a high degree of asymptomatic Omicron infection. However, a corresponding increase in the spike antibody titers was not observed. It is likely that spike antibodies may have waned faster than nucleocapsid antibodies between weeks 25 and 40 or that Omicron-specific spike antibodies elicited during the wave were not detected using the spike protein against the WT strain.

In the KEMHRC cohort, the Omicron wave hit between weeks 11 and 15. Parallelly, the anti-nucleocapsid titers increased between weeks 12, 14 and 25. At the post-Omicron sampling point at week 25, spike antibody levels were similar between Covaxin^®^ and Covishield™ arms in both the serogroups. Between weeks 25 and week 40, the decline in spike antibody titers were fairly low and additional sampling will be needed to address comparison between vaccines.

In the SUHRC cohort, the Omicron wave occurred between weeks 11 and 15. Preliminary results suggest that the trends were similar to the KEMHRC cohort that was exposed around similar time frames.

In the SJRI cohort, the Omicron wave hit during the primary vaccination period and manifested as higher post-vaccination spike as well as nucleocapsid antibody titers than other cohorts in the Covaxin^®^ arm. In both seronegative individuals and seropositive individuals, antibody decline by week 25 appeared to be faster in the Covaxin^®^ arm than in the Covishield™ arm. However, a direct comparison of the vaccine-specific antibody decay rates could not be made due to the differences in the time between vaccination, sampling and infection.

### Breadth of Antibody Responses Against SARS-CoV-2 Variants of Concern (VoCs)

We next used the ACE2 inhibition assay on the MSD platform to measure surrogate neutralization against the major SARS-CoV-2 lineages and sub-lineages at all timepoints.

In seronegative individuals, vaccination with Covishield™ consistently elicited higher ACE2 inhibition activity than Covaxin^®^ against Wild-type (WT) and Delta spikes in both the BBH and KEMHRC cohorts (Figure 4C; Table 4). Neither vaccine elicited Omicron-specific antibodies. The Omicron wave was associated with increased activity against WT, Delta and BA.1 in the BBH cohort at weeks 25-40, though vaccine-specific differences cannot be assessed due to differences in the timing of exposure and sampling. The Omicron wave elicited higher responses against WT and Delta than against BA.1. Similar trends were observed in the KEMHRC cohort that was exposed to the Omicron at a much earlier time post-vaccination than the BBH cohort. Interestingly, in the SJRI cohort, where participants were co-exposed to WT strain by vaccination and the Omicron strain through infection, stronger responses were elicited against WT than against Omicron. Unlike the sparse sampling in the BBH and KEMHRC cohorts, the SJRI cohort was sampled very closely during the Omicron wave, but still did not detect strong Omicron responses. This suggests that the Omicron spike in asymptomatic infection may not be as immunogenic as WT, though this needs to be confirmed by measurement of mucosal immune responses.

**Table 4.** Inhibition of ACE2 binding to SARS-CoV-2 WT, Delta AY.4 and Omicron BA.1 spike in all cohorts across all study timepoints. Data are shown as N and median ^36^

| Week<br>s | BBH |  |  |  | KEMHRC |  |  |  | SUHRC |  |  |  | SJRI |  |  |  |
| --- | --- | --- | --- | --- | --- | --- | --- | --- | --- | --- | --- | --- | --- | --- | --- | --- |
|  | Covaxin |  | Covishield |  | Covaxin |  | Covishield |  | Covaxin |  | Covishield |  | Covaxin |  | Covishield |  |
|  | N | Median | N | Median | N | Median | N | Median | N | Median | N | Median | N | Median | N | Median |
| **Seronegative** |  |  |  |  |  |  |  |  |  |  |  |  |  |  |  |  |
| **WT Spike, % inhibition** |  |  |  |  |  |  |  |  |  |  |  |  |  |  |  |  |
| 0 | 20 | 4.2 | 46 | 1.6 | 20 | 1.5 | 19 | 4.1 | 4 | 2 | 7 | 4.6 | 7 | 3.1 | 12 | 8.5 |
| 4 | 20 | 0.0 | 46 | 21.4 | 20 | 5.6 | 19 | 21.8 | 4 | 1 | 7 | 14.7 | 7 | 11.2 | 12 | 31.6 |
| 6 | 20 | 12.0 |  |  | 20 | 18.9 |  |  | 4 | 15.7 |  |  | 7 | 17.9 |  |  |
| 12 | 20 | 4.0 | 46 | 8.4 | 20 | 11.2 | 19 | 20.5 | 4 | 67.9 | 7 | 72.7 | 7 | 12.2 | 12 | 85.7 |
| 14 |  |  | 46 | 60.2 |  |  | 19 | 65.0 |  |  | 7 | 92.4 |  |  | 12 | 79.7 |
| 25 | 20 | 3.7 | 43 | 28.8 | 20 | 44.9 | 19 | 67.0 | 3 | 19.3 | 7 | 65.3 | 6 | 5.9 | 12 | 51.6 |
| 40 | 17 | 30.2 | 37 | 77.5 | 15 | 26.7 | 15 | 39.7 | 4 | 57.9 | 7 | 82.2 |  |  |  |  |
| **Delta AY.4 Spike, % inhibition** |  |  |  |  |  |  |  |  |  |  |  |  |  |  |  |  |
| 0 | 20 | 1.0 | 46 | 0.0 | 20 | 0.0 | 19 | 2.5 | 4 | 0 | 7 | 0.9 | 7 | 0.0 | 12 | 3.4 |
| 4 | 20 | 0.0 | 46 | 11.9 | 20 | 5.1 | 19 | 12.8 | 4 | 0 | 7 | 12.3 | 7 | 8.4 | 12 | 17.5 |
| 6 | 20 | 10.4 |  |  | 20 | 8.1 |  |  | 4 | 2.6 |  |  | 7 | 11.4 |  |  |
| 12 | 20 | 0.0 | 46 | 5.3 | 20 | 9.9 | 19 | 9.3 | 4 | 54.1 | 7 | 51.1 | 7 | 9.6 | 12 | 63.6 |
| 14 |  |  | 46 | 45.0 |  |  | 19 | 48.3 |  |  | 7 | 78.7 |  |  | 12 | 58.2 |
| 25 | 20 | 0.4 | 43 | 16.8 | 20 | 34.4 | 19 | 47.2 | 3 | 14.1 | 7 | 61.4 | 6 | 1.7 | 12 | 33.8 |
| 40 | 17 | 26.9 | 37 | 60.6 | 15 | 21.3 | 15 | 32.1 | 4 | 49.5 | 7 | 68 |  |  |  |  |
| **Omicron BA.1 Spike, % inhibition** |  |  |  |  |  |  |  |  |  |  |  |  |  |  |  |  |
| 0 | 20 | 0.0 | 46 | 0.0 | 20 | 2.3 | 19 | 0.0 | 4 | 0 | 7 | 11.7 | 7 | 0.0 | 12 | 8.6 |
| 4 | 20 | 0.0 | 46 | 0.0 | 20 | 0.0 | 19 | 0.0 | 4 | 0 | 7 | 0 | 7 | 0.0 | 12 | 0.0 |
| 6 | 20 | 0.0 |  |  | 20 | 0.0 |  |  | 4 | 0 |  |  | 7 | 0.0 |  |  |
| 12 | 20 | 0.0 | 46 | 0.0 | 20 | 0.0 | 19 | 0.0 | 4 | 11.1 | 7 | 6.3 | 7 | 0.0 | 12 | 19.0 |
| 14 |  |  | 46 | 0.0 |  |  | 19 | 0.0 |  |  | 7 | 17.6 |  |  | 12 | 17.3 |
| 25 | 20 | 0.0 | 43 | 0.0 | 20 | 11.5 | 19 | 0.0 | 3 | 0 | 7 | 10.8 | 6 | 0.0 | 12 | 9.7 |
| 40 | 17 | 0.0 | 37 | 18.5 | 15 | 4.8 | 15 | 0.0 | 4 | 3.1 | 7 | 20.1 |  |  |  |  |
| **Seropositives** |  |  |  |  |  |  |  |  |  |  |  |  |  |  |  |  |
| **WT Spike, % inhibition** |  |  |  |  |  |  |  |  |  |  |  |  |  |  |  |  |
| 0 | 48 | 23.2 | 61 | 19.0 | 47 | 18.5 | 29 | 13.4 | 9 | 10 | 24 | 10.7 | 29 | 14.3 | 24 | 10.4 |
| 4 | 48 | 33.8 | 61 | 99.0 | 47 | 32.9 | 29 | 99.1 | 9 | 23.2 | 24 | 99.1 | 29 | 72.3 | 24 | 99.1 |
| 6 | 48 | 39.0 |  |  | 47 | 47.8 |  |  | 9 | 38.7 |  |  | 29 | 60.5 |  |  |
| 12 | 47 | 27.3 | 61 | 86.4 | 47 | 41.6 | 29 | 85.9 | 9 | 34.8 | 24 | 95.7 | 29 | 57.8 | 24 | 98.2 |
| 14 |  |  | 61 | 92.4 |  |  | 29 | 95.4 |  |  | 24 | 96.1 |  |  | 24 | 97.9 |
| 25 | 43 | 16.0 | 61 | 70.6 | 47 | 66.0 | 29 | 82.5 | 8 | 53.7 | 24 | 83.1 | 26 | 36.5 | 24 | 92.9 |
| 40 | 29 | 46.1 | 55 | 80.9 | 30 | 46.2 | 24 | 80.8 | 9 | 50.4 | 24 | 84 |  |  |  |  |
| **Delta AY.4 Spike, % inhibition** |  |  |  |  |  |  |  |  |  |  |  |  |  |  |  |  |
| 0 | 48 | 24.9 | 61 | 21.7 | 47 | 13.6 | 29 | 11.6 | 9 | 12.7 | 24 | 11.1 | 29 | 12.6 | 24 | 7.5 |
| 4 | 48 | 38.3 | 61 | 98.6 | 47 | 29.8 | 29 | 98.2 | 9 | 21.1 | 24 | 98.6 | 29 | 77.4 | 24 | 97.8 |
| 6 | 48 | 42.0 |  |  | 47 | 48.2 |  |  | 9 | 24.4 |  |  | 29 | 59.1 |  |  |
| 12 | 47 | 33.5 | 61 | 83.6 | 47 | 47.8 | 29 | 85.3 | 9 | 30.3 | 24 | 94.4 | 29 | 55.2 | 24 | 98.0 |
| 14 |  |  | 61 | 89.6 |  |  | 29 | 92.0 |  |  | 24 | 95.1 |  |  | 24 | 97.3 |
| 25 | 43 | 22.1 | 61 | 62.0 | 47 | 63.5 | 29 | 70.9 | 8 | 49.1 | 24 | 83.1 | 26 | 39.8 | 24 | 89.5 |
| 40 | 29 | 60.8 | 55 | 76.8 | 30 | 45.6 | 24 | 75.4 | 9 | 66 | 24 | 80 |  |  |  |  |
| **Omicron BA.1 Spike, % inhibition** |  |  |  |  |  |  |  |  |  |  |  |  |  |  |  |  |
| 0 | 48 | 0.0 | 61 | 0.0 | 47 | 0.0 | 29 | 0.0 | 9 | 0 | 24 | 0 | 29 | 0.0 | 24 | 0.0 |
| 4 | 48 | 0.0 | 61 | 41.4 | 47 | 0.0 | 29 | 43.7 | 9 | 0 | 24 | 43.1 | 29 | 23.7 | 24 | 62.7 |
| 6 | 48 | 0.0 |  |  | 47 | 0.0 |  |  | 9 | 0 |  |  | 29 | 14.6 |  |  |
| 12 | 47 | 0.0 | 61 | 0.0 | 47 | 0.0 | 29 | 17.0 | 9 | 0 | 24 | 13.4 | 29 | 11.9 | 24 | 55.7 |
| 14 |  |  | 61 | 11.4 |  |  | 29 | 19.8 |  |  | 24 | 29.7 |  |  | 24 | 54.7 |
| 25 | 43 | 0.0 | 61 | 9.8 | 47 | 14.5 | 29 | 11.9 | 8 | 6.6 | 24 | 15.2 | 26 | 4.5 | 24 | 27.4 |
| 40 | 29 | 17.9 | 55 | 22.1 | 30 | 7.8 | 24 | 13.8 | 9 | 6.1 | 24 | 1.4 |  |  |  |  |

In seropositive individuals, Covishield™ vaccination elicited strong and near-saturation levels of antibodies against both WT and Delta spikes in all three cohorts (Figure 4D; Table 4). Interestingly, a single dose of Covishield™ elicited relatively strong Omicron-specific responses in seropositive individuals. However, a second dose of Covishield™ did not increase responses and the median activity remained lower than after the first dose. Vaccination with Covaxin^®^ increased activity against WT and Delta but not against Omicron. At week 4, Omicron-specific responses were higher in the infection plus vaccination SJRI cohort than in the breakthrough-infection cohorts of BBH and KEMHRC. Correspondingly, Omicron-specific responses waned slower in the SJRI cohort than in the BBH and KEMHRC cohorts.

Overall, the breadth and potency of responses against SARS-CoV-2 VoCs was higher with Covishield™ than with Covaxin^®^, and in seropositive individuals than in seronegative individuals.

## Discussion

In a longitudinal study that includes pre-vaccination baselines, we compared the immune responses elicited by the two main SARS-CoV-2 vaccines utilized in India. The comparison was designed to explore vaccination-induced immune responses in both seronegative and seropositive participants. In general, the data show that Covishield™ elicited higher magnitude immune responses than Covaxin^®^. This is supported by 4 lines of evidence as described below. Firstly, in seronegative participants, the adenovirus vectored vaccine, Covishield™, elicited near-complete seroconversion rates between and 100%. Seroconversion for Covaxin^®^ was more variable, reaching > 90% only in cohorts that were vaccinated and therefore had samples collected closer to the Omicron wave (the SUHRC and SJRI cohorts). Seroconversion for Covaxin^®^ in the pre-Omicron cohorts (52.0 and 75.7% for BBH and KEMHRC cohorts, respectively) was much lower than rates reported from the phase 2 trial^13^. This difference may be due to the method of testing for antibodies. Our study measured seroconversion using a binding antibody assay based on stabilized spike trimer protein (DiaSorin TrimericS). This assay is reported to be selective for neutralizing rather than non-neutralizing antibodies whereas the phase 2 trial utilized in-house assays which have not been validated.

Similar to seronegative individuals, seroconversion in seropositive individuals was higher in Covishield™ than in Covaxin^®^ recipients. A significant proportion of seropositive Covaxin^®^ recipients (37.4%) did not respond to vaccination.

Secondly, in addition to greater rates of seroconversion, Covishield™ also elicited higher antibody titers (GMT) than Covaxin^®^ in both seronegative individuals and seropositive individuals. These trends were observed in three cohorts that had completed vaccination prior to the Omicron wave. The fourth cohort, in which the vaccination overlapped with the Omicron wave, showed comparable post-vaccination antibody titers in the Covishield™ and Covaxin^®^ recipients. However, antibody titers in the fourth cohort with delayed recruitment confirmed that the waning of the immune response was slower in Covishield™ recipients. Importantly, we also confirmed these findings using an independent assay platform, the MSD platform which has a higher dynamic range.

Thirdly and in parallel to the antibody responses, in the earliest cohort (BBH), Covishield™ elicited a higher frequency of both CD4 and CD8 T cells while Covaxin^®^ elicited mainly CD4 T cell responses. The frequencies and quality of cytokine-expressing T cells that we report in the seronegative Covishield™ arm are broadly consistent with prior studies, though the dosing interval in those studies was only 4 weeks^14,15^. The poor CD8 responses from Covaxin^®^ is not unexpected given that TLR7/8 agonists need to be physically linked to the antigen in order to induce CD8 T cell responses in mice^16^, which is not the case with Covaxin^®^. In humans, however, *in vivo* antigen synthesis using either live viruses, non-replicating virus vectors or mRNA/DNA vaccines and classical MHC-I-presentation of antigen remain the only ways to elicit robust anti-viral CD8 T cells. Nevertheless, vaccine-elicited CD8 T cell responses in the blood have not yet been demonstrated as a correlate of protection (CoP) against severe COVID-19 in humans, and therefore the significance of spike-specific CD8 T cell responses is not clear. In seropositive participants, the vectored vaccine elicited higher rates of seroconversion and antibody magnitude without a strong expansion of pre-existing spike-specific CD4 T cells in the blood. In both vaccine groups, lack of immunogenicity in a subset of seropositive individuals was associated with higher pre-vaccination antibody titers. This is consistent with studies on mRNA vaccines in seropositive subjects^17,18^. The negative role of high levels of circulating antibodies on vaccine immunogenicity and/or efficacy has also been observed with other vaccines^19–21^. The high levels of anti-SARS-CoV-2-spike seropositivity in the population suggests that the development of spike-based pan-coronavirus vaccines and their deployment through public health campaigns must consider the immune history of the population.

Lastly, in a subset of participants, we tested neutralizing responses using an authentic WT virus assay and surrogate neutralizing responses using an ACE-2 inhibition assay against several SARS-CoV-2 variants. In every virus variant tested, Covishield™ elicited higher responses than Covaxin^®^.

The superiority of vectored vaccines over inactivated vaccines for antibody responses against SARS-CoV-2 has been demonstrated by other studies^7–11^. Further, inactivated viral vaccines have historically been of lower immunogenicity, requiring 4 doses in the primary series for the inactivated polio vaccine and the rabies vaccine. The inactivated Hepatitis A vaccine is given as two doses, but the doses are spaced six months apart. In contrast, the primary immunization series of all inactivated Covid vaccines have been only two doses spaced 2-4 weeks apart. However, as a sole exception, the inactivated vaccine VLA-2001 (Valneva’s inactivated COVID-19 vaccine manufactured using the Vero cell platform) demonstrated non-inferiority for seroconversion rate and superiority for neutralizing titers against ChAdOx1-S^22^ (developed by Oxford-AstraZeneca and manufactured by AstraZeneca; same as Covishield™ tested in this study).

Notwithstanding the lower magnitude of spike antibodies reported in this study, Covaxin^®^ has demonstrated comparable or only moderately lower efficacy against SARS-CoV-2 variant waves from ancestral to Delta^23,24^, suggesting that non-neutralizing spike-binding antibodies and antibodies against non-spike virion proteins could be contributors to the correlates of protection (CoP). All available vectored and mRNA-based Covid vaccines are based on the spike protein and therefore published CoP studies from the developed world have only been able to address the role of antibodies against the spike protein^25–28^.

Emerging pre-clinical data suggest a potential role for vaccine-elicited nucleocapsid responses in controlling virus replication in the lung^29^. Though the limited sequence evolution in nucleocapsid compared to spike could make nucleocapsid an attractive target, the shorter durability of circulating antibodies against nucleocapsid compared to spike following infection necessitates the study of the comparative durability of vaccine-elicited nucleocapsid versus spike antibodies. Here, we show that Covaxin^®^ elicits nucleocapsid antibodies but that those levels seem to be sub-optimal given that levels increase further upon breakthrough Omicron infection. It is likely that nucleocapsid antibodies and potential Fc-dependent antibody effector functions elicited by Covaxin^®^ lower the threshold of anti-spike neutralizing antibodies required for protection. In a study which considered neutralizing titres to predict protective efficacy, early phase clinical trial data from Covaxin^®^ was included. Modelling in this study suggests that Covaxin^®^ might be as good, if not better, than Covishield™ in efficacy ^30^. However, our data seems to suggest otherwise. The lack of planned (or publicly accessible) CoP analyses for Covaxin^®^ are a missed opportunity for a more coordinated global vaccine development response.

As has been shown by others in seronegative individuals, two doses of vaccine did not elicit Omicron-specific responses^31–34^. Interestingly, in seropositive individuals, only a single dose of Covishield™ elicited clearly measurable Omicron-specific responses assayed using inhibition of ACE2 binding. Most other studies that demonstrated the need for three or four exposures to spike before elicitation of Omicron-specific neutralizing responses were unable to study the effect of second exposure due to cohort sampling limitations. It remains possible that prior exposure to Delta rather than ancestral strain may have led to better Omicron responses in the seropositive cohort in this study. Curiously, a second dose of Covishield™ did not elicit the same level of anti-Omicron activity as the first dose and needs further investigation.

The cohort that we have studied here is unique in being first exposed to the Delta variant through infection followed by exposure to the ancestral strain through vaccination, while other studies were predominantly based on ancestral-strain based vaccination or infection followed by breakthrough infection with variants. The exposure to Delta prior to the ancestral sequence may have pre-disposed the immune system to have broader responses and can be addressed further by measuring variant-specific T cell responses. In summary, we show that a vectored vaccine elicits stronger and broader anti-spike responses than an inactivated vaccine. We also show that Omicron-specific antibody responses from asymptomatic infection wane faster than WT- and Delta-specific antibody responses. Non-responsiveness to vaccination in seropositive subjects is associated with higher baseline antibody titers. The role of the duration between the Delta wave and vaccination in driving both antibody and cellular immune responses is under investigation.

It remains to be seen how the withdrawal of the zero-COVID policy will affect the largely uninfected Chinese population which was either vaccinated with the original ancestral strain based inactivated virus vaccine (Sinovac or Sinopharm) a year or more ago or remained unvaccinated, given that the currently circulating Omicron variants have diverged significantly from the ancestral viruses used for making these vaccines. Public health programmes which are making plans now for expanding booster dose coverage will benefit from the use of heterologous boosters rather than repeating the homologous primary dose. This has been demonstrated by studies that examined homologous and heterologous boosting with Covaxin^®^ and Covishield^TM35^, Coronavac followed by BBIBP-CorV, AZD1222 or BNT162b2^36^ and AZD1222 followed by AZD1222, NVX-CoV2373, VLA2001, BNT162b2, mRNA-1273, Ad26.CoV2.S or CVnCoV^5^. These findings have implications for booster vaccine policy-making. They suggest that public health vaccination campaigns should be followed-up with extensive immunological analyses of the general population using validated assays. These findings also indicate that the development and deployment of booster doses and pan-coronavirus vaccines for populations with high levels of exposure and vaccine coverage should take into account the prevailing magnitude and quality of immune responses in the population. Further, CoP analyses must be mandatory and planned in advance for vaccines administered through the national health programme.

### Study Limitations

In this study, we have not specifically measured the levels of non-neutralizing antibodies or non-neutralizing functions that may be important for vaccine efficacy, and the durability of the vaccine-elicited immune responses could not be assessed due to the Omicron wave. Asymptomatic Omicron infection referred to in this study is solely based on antibody titers measured several weeks apart and does not have a concomitant RT-PCR test. The sparce sampling during the Omicron wave in most of the participants also limits direct quantitative comparisons of the vaccines during the Omicron wave. While it appears that Covaxin^®^ vaccination was associated with poor Omicron responses during breakthrough asymptomatic infection and was likely due to prior imprinting from non-neutralizing antibodies or narrow WT-specific neutralizing antibodies, further studies will be needed to assess this phenomenon. Our study population is limited to pre-vaccination serostatus and not to confirmed prior infection or exposure. It therefore remains possible that while most seronegative participants may have remained unexposed between the beginning of the pandemic and the time of study recruitment, a subset of seronegative individuals may have been exposed previously but seroreverted at the time of recruitment due to decline in circulating antibody titers. In the absence of a CoP for Covaxin^®^, the immunogenicity results cannot be extrapolated to infer vaccine efficacy.

## Methods

### Study design and participants

This study was done in healthy adults aged 18-45 years at Bangalore Baptist Hospital (BBH), National Centre for Biological Sciences (NCBS), Institute for Stem Cell Science and Regenerative Medicine (InStem), King Edward Memorial Hospital Research Center (KEM), Symbiosis University Hospital and Research Center (SUHRC), National Chemical Laboratory (NCL), Indian Institute of Science Education and Research Pune (IISER-Pune), and St. John’s Research Institute (SJRI). The trial was approved by the Institutional Ethics Committee (BBH/IRB/2021/33, NCBS/IEC-27/01, inStem/IEC-23/02, IISERP-IHEC/Admin/2021/018, SIU/IEC/298, KEMHRC/RVM/EC/1225, NCL-IHEC/SEP2021/001, SJRI-298/2021) at each site and registered at the Clinical Trial Registry of India (CTRI) website (CTRI/2021/09/036258).

SARS-CoV-2 vaccine-naïve participants were screened for serostatus and recruited at BBH, KEMHRC, SUHRC and SJRI using a combination of anti-spike and/or anti-nucleocapsid antibodies, either qualitative or quantitative, manufactured by either Roche Diagnostics, Abbott Laboratories or Liaison DiaSorin at each clinical site. Recruitment was done using a combination of unbiased as well serostatus-confirmed inclusion. Baseline samples were re-tested and classified for their serostatus using DiaSorin TrimericS and MSD platforms at Christian Medical College (CMC), Vellore. Sample size calculations were not performed and recruitment numbers were based on the capacity at each site. All the participants were enrolled after obtaining informed written consent. Participants with a history of medical illness or prior severe COVID-19 that required ventilation or administration of biologics such as convalescent plasma or monoclonal antibodies were excluded. Vaccine-naïve participants were administered two doses of either Covaxin^®^ at 4-week interval or Covishield™ at 12-week interval per the prevailing government norms. Allocation of participants to vaccine arms was non-randomized and per participant-choice. Participants in the Covaxin^®^ arm were sampled at Day 0 (prior to first dose of vaccine), Day 28 (prior to second dose of vaccine), Day 42, Day 84, month 6 and month 9. Participants in the Covishield™ arm were sampled at Day 0 (prior to the vaccine dose), Day 28, Day 84 (prior to the second dose), Day 98, month 6 and month 9. Participants were followed bi-monthly for symptoms of COVID-like illness.

### Procedures

Binding antibodies against the trimeric spike protein in all study participants were measured using the LIAISON SARS-CoV-2 TrimericS IgG assay on the DiaSorin platform per the manufacturer’s instructions. The assay cut-off of 33.8 BAU/ml was used to determine seropositivity. In samples with titer <33.8 BAU/ml, seroconversion was defined as an increase in titer to >33.8 BAU/ml following vaccination. In samples with titer >33.8 BAU/ml, seroconversion was defined as atleast 2-fold increase in titer following vaccination.

Binding antibodies against trimeric spike protein of SARS CoV-2, nucleocapsid, receptor binding domain (RBD) and N-terminal domain (NTD) were measured using the MSD V-Plex SARS-CoV-2 Panel 1 (IgG) kit. ACE-2 inhibition activity was measured using the MSD V-Plex SARS-CoV-2 Panel 13 (ACE2) and Panel 25 (ACE2) kits at a 1:50 dilution per the manufacturer’s instructions.

Neutralizing antibodies were measured in a micro-neutralization assay. This assay was performed with institutional biosafety clearances TFR:NCBS:37IBSC2021/RP, inStem/G-141(3)-20/CJ, inStem/G-141(3)-20/AK and NCBS:37IBSC2021/VS1. Vero E6 cells were seeded at 10,000 cells per well in a 96-well tissue culture plate. After 24 hours, heat inactivated plasma samples were diluted in DMEM supplemented with 2% FBS in a two-fold dilution series. 75 μL of diluted plasma was mixed with 75 μL of SARS-CoV-2 Italian INMI1 strain (NR-52284, BEI resources; MOI=0.01). This mixture was incubated for 1 hour in a humidified 37ºC incubator supplemented with 5% CO_2_. The plasma-virus mixture was then added to the cell monolayer and incubated for 1 hour in the incubator. The plasma-virus mixture was removed from the cell monolayer, and the cells are replenished with fresh media supplemented with 2% FBS and incubated for 48hrs. Duplicate wells containing cells only and cells with virus diluent were included as controls. A previously titered plasma sample was used as positive control in all the assays. After 48 h of incubation. Microscopic images are recorded using an inverted optical microscope and Q-capture pro-7.0 software. Neutralization titer was defined as the highest dilution of plasma at which there was complete rescue of virus-induced cytopathic effects on the cell monolayer. Representative images are shown in Supplementary Figure 3A.

T cell responses were measured using frozen PBMC (isolated from heparinized blood) specimens per standard protocols. Briefly, cells were thawed and rested for 4 hours. 1.5 million cells/well were stimulated with 1 μg/ml of SARS-CoV-2 Prot S Complete, or a mix of Prot N and Prot M at 1 μg/ml each in a 37 ºC/5% CO_2_ incubator. Cells left unstimulated or stimulated with PHA-P served as negative and positive controls for every sample. Co-stimulatory antibodies (1 μg/ml each of anti-CD28 and anti-CD49d) were included in all test conditions. After 2h, Brefeldin A was added to all wells for a final concentration of 5 μg/ml. Cells were incubated for an additional 16 h. At the end of the stimulation period, cells were washed and stained with viability dye for 10 min at room temperature, followed by incubation with a cocktail of surface stain antibodies (anti-CD14 BV650, anti-CD16 BV650, anti-CD20 BV650, anti-CD4 Alexa700 and anti-CD8 BV605) for 20 min at 4 ºC. After washing, cells were fixed, permeabilized and stained with a cocktail of intracellular stain antibodies (anti-CD3 Cy7APC, anti-IFNγ BV421, anti-IL2 PE and anti-TNF BV785) for 20 min at 4 ºC. After additional washes, cells were fixed again and acquired on an LSR Fortessa within 24h. Data were analyzed using FlowJo 10.8.1. A control sample treated in the same way as the samples was included in every run of the assay. Paired samples from the same participant were tested within the same run. Cytokine gates were set using the unstimulated controls and maintained for a given run. A representative gating strategy is shown in Supplementary Figure 4A. Antigen-specific frequencies were calculated by subtraction of background responses from the unstimulated control of each sample. Assay limit of detection was set to 0.002% based on inclusion criteria of acquiring minimum 50,000 live CD4+ T cells in each sample. Responses less than 0.002% were set to 0.002%.

### Statistical Analysis

For antibody analyses, within-group comparisons across timepoints were tested using two-way ANOVA with Tukey’s correction. Between-group comparisons after each vaccine dose were tested using two-way ANOVA with Sidak’s correction. Differences between pre- and post-vaccination T cell responses were compared using Kruskal-Wallis test with Dunn’s correction. All statistical analyses were performed with GraphPad Prism 9.4.1.

## Data Availability

All data produced in the present study are available upon reasonable request to the authors

## Author contributions

**Conceptualization and Study Design:**MA, SB, GD, BR, DA, CE, JJ, AK, MD, AB, PR, SLS, AV, VB, GK, SM

**Data Curation:** MA, SB

**Formal Analysis:** MA

**Funding Acquisition:** SLS, GK, SM

**Investigation:** SB, PN, VS, AP, SI

**Project Administration:** GD, AP, PS, RG, BR, DA, PS, MA, SS, UD, CB, MJ, SC, AG, CJ, AJ, NR, GD, VR, CE, AK, MD, AB, PR, SM

**Resources:** RFJ, PN, VS, AP, VM, JB, KP, AT, GM, RG, BR, SR, DA, SS, AS, PP, RS, SG, AU, PA, AN, ST, MB, RY, MV, SS, UD, CB, MJ, LRI, AG, CJ, AK, VS, AJ, NR, SR, VA, GD, VR, CE, AK, MD, AB

**Supervision:** MA, CJ, CE, AK, AC, AR, MD, AB, PR, SLS, AV, VB, GK, SM

**Visualization:** MA, SM

**Writing-Original draft preparation:** MA, SM

**Writing-Review and Editing:** All authors

## Acknowledgements

The project was fully supported by CSR funding from Hindustan Unilever Limited (HUL) and Unilever India Pvt. Ltd. (UIPL) and facilitated by the Office of the Principal Scientific Advisor (Government of India). We acknowledge Dr. Vibhav Sanzgiri and Dr. Janhavi Raut (HUL/UIPL), Dr. Sapna Poti (PSA Office) and Dilip Chenoy (former Secretary General FICCI and DC Skills) for their encouragement and support. Department of Atomic Energy (Government of India) under Project No. RTI 4006 to NCBS, and the facilities provided by the Bangalore Life Science (BLiSc): Biosafety-I, II, and III, Central Imaging and Flow Facility (CIFF), Instrumentation, Research Development Office, are gratefully acknowledged. Dr. Vineetha Raghavan and Knight Paul Pandian at NCBS for administrative support, Saurabh Vaishnav for training and access to the flow cytometry facility at IISER Pune, Dr. Rama Akondy at Ashoka University for advice on T cell assays, and the use of facilities at NCL are also gratefully acknowledged. The following reagent was deposited by Dr. Maria R. Capobianchi for distribution through BEI Resources, NIAID, NIH: SARS-Related Coronavirus 2, Isolate Italy-INMI1, NR-52284. SM is supported by a J.C. Bose Fellowship from the Department of Science and Technology and is a Margadarshi Fellow of the DBT-Wellcome Trust India Alliance (IA/M/15/1/502018).

## Figure Legends

**Figure 1 Participant recruitment and vaccination schedule in the context of Delta and Omicron waves and vaccination status in the country**.A) Recruitment schedules across the sites. Horizontal colored lines show the time span during which the first and second doses of vaccine were administered in each site. Solid lines represent the first and second dose of Covaxin^®^ and dotted lines represent the first and second dose of Covishield. B) Participant recruitment and follow-up in each of the cohorts.

**Figure 2 Antibody responses following vaccination in the per-protocol population from all cohorts**

Antibody responses against ancestral spike were measured on the Diasorin TrimericS platform in the per-protocol population across the full cohort. Participants were classified as either seronegative (n=82 for Covaxin^®^, open red circles; 117 for Covishield, open blue squares) or seropositive (n = 139 for Covaxin^®^, filled red circles; 168 for Covishield, filled blue squares) at baseline using the assay cut-off of 33.8 BAU/ml (horizontal dotted line). Data are shown as geometric mean ± 95% CI. Within-group comparisons to the baseline were assessed using a two-way ANOVA with Tukey’s correction for multiple comparisons. B) Bar graphs show seroconversion rates. Seroconversion was defined as change from <33.8 to >33.8 for those with titers <33.8 BAU/ml prior to the vaccine dose. For those with titers >33.8 BAU/ml, seroconversion was defined as increase in titer by atleast 2-fold following the vaccine administration. C) Between group comparisons after each vaccine dose were made using two-ANOVA with Sidak’s correction. D) Neutralizing responses were measured as inhibition of cytopathic effects by Italian strain INMI1 on VeroE6 cells in a microneutralization assay (see Supp fig. 3A). Participants were classified as either seronegative or positive using a cut-off of 33.8 BAU/ml for ancestral spike binding on the TrimericS platform. Samples from baseline and 2-weeks post the second dose of vaccine were tested. Their corresponding antibody trajectories are shown in Supp figs. 3C and D. Statistical comparisons were made using two-way ANOVA with Sidak’s correction. For all statistical analyses, p-values >0.05 are shown as ns (not significant).

**Figure 3 Vaccine-induced T cell responses** Frequency of spike-specific CD4 or CD8 T cells expressing at least one cytokine amongst IFNγ, IL2 and TNF in response to stimulation with either spike peptide pool, mixture of nucleocapsid and matrix peptide pools or PHA, following vaccination in A) Seronegative Covaxin^®^ (red) or Covishield (blue) Seropositive Covaxin^®^ (red) or Covishield (blue). Frequencies are shown after background subtraction from the unstimulated control. Differences between pre-and post-vaccination responses were compared using Kruskal-Wallis test with Dunn’s correction. p-values >0.05 are shown as ns (not significant).

**Figure 4 Durability and breadth of antibody responses following vaccination and exposure to the Omicron wave**. A, B) Durability of antibody responses against spike protein (both Diasorin TrimericS and MSD platforms) and nucleocapsid protein (MSD) are shown for each cohort as geometric mean ± 95% CI for seronegative individuals (A) and seropositive individuals (B). C, D) Inhibition of ACE2 binding to the indicated spike proteins is shown as medians with interquartile range for seronegative individuals (C) and seropositive individuals (D). Trajectories for individual participants are shown as thin background lines. Horizontal bars (red - Covaxin^®^, blue - Covishield) within each graph show timepoints in which sampling was performed during the Omicron wave

**Supplementary figure 1 Classification of study participants based on antibody trajectories following vaccination** A) Seronegative individuals B) Seropositive individuals. Arrowheads indicate the time of vaccination.

**Supplementary Figure 2 Change in antibody titers within each study group across cohorts**. Antibody responses against ancestral spike measured on the Diasorin TrimericS platform for per-protocol population across each site. Participants were classified as either seronegative or seropositive at baseline using the assay cut-off of 33.8 BAU/ml (horizontal dotted line). Stars within each graph indicate timepoints where sampling in greater than 25% of the population from the cohort was performed during the Omicron wave in January and February 2022. Colors of the stars indicate the cohort. Statistical comparisons between cohorts at each timepoint were performed using a two-way ANOVA with Tukey’s correction. B) summary of p-values

**Supplementary figure 3** A) Representative light microscope images showing determination of neutralizing titers. The highlighted image (1:40) shows the highest dilution of plasma at which there is complete rescue of virus-induced cytopathic effects. B) Pearson correlation between neutralization titers against authentic virus and either anti-spike binding IgG or percent inhibition of ACE-2 binding to ancestral spike protein. B,C) Binding antibody titers in participants who were included for neutralization assay from seronegative and seropositive groups respectively.

**Supplementary figure 4** A) Representative gating strategy for flow cytometry-based quantification of antigen-specific T cells expressing IFNγ, IL2 and/or TNF. B,C) Antibody responses against spike and nucleocapsid proteins in the seronegative and seropositive participants included in the ICS assay.

**Supplementary Figure 5 T cell responses prior to vaccination in seronegative versus seropositive participants**. A) T cell responses against a pool of peptides derived from either ancestral spike or a mixture N and M or control PHA measured by ICS in one of the cohorts. Background responses from an unstimulated control were subtracted. Participants were classified as either seronegative (n=25) or seropositive (n=44) against SARS-CoV-2 spike at baseline using the MSD assay. Solid horizontal line (red) indicates the group median. Differences between seronegative individuals and seropositive individuals were assessed using Kruskal-Wallis test with Dunn’s correction. B) Antigen-specific responses are shown for each of the indicated cytokines. Bar heights represent the group median. Differences between seronegative individuals and seropositive individuals were assessed using Kruskal-Wallis test with Dunn’s correction. C) Pearson correlation between antibody titers and spike-specific or nucleocapsid-specific CD4 T cells prior to vaccination

**Supplementary Figure 6**

A, B) Spike-specific T-cell responses among participants classified as either responders or non-responders based on antibody titers measured onA) TrimericS or B) MSD platform. Responders were classified as those whose post-vaccination antibody titers were greater than either 33.8 BAU/ml on Trimeric S or 1960 AU/ml (MSD units). Statistical comparisons were made using Kruskal-Wallis test with Dunn’s correction. Bar heights represent group median C) T-cell responses shown against individual cytokines pre- and post-vaccination against the indicated antigens (either spike, mixture of N and M or PHA). Statistical comparisons were made using Kruskal-Wallis test with Dunn’s correction. Bar heights represent group median

**Supplementary Figure 7**

A, B) Spike-specific T-cell responses among participants classified as either responders or non-responders based on antibody titers measured on A) TrimericS or B) MSD platform. Responders were classified as those whose post-vaccination antibody titers were atleast 2-fold greater than at baseline. Statistical comparisons were made using Kruskal-Wallis test with Dunn’s correction. Bar heights represent group median C) T-cell responses shown against individual cytokines pre- and post-vaccination against the indicated antigens (either spike, mixture of N and M or PHA). Statistical comparisons were made using Kruskal-Wallis test with Dunn’s correction. Bar heights represent group median

