## Supplemental Figures for "Interim results from comparison of immune responses elicited by an inactivated and a vectored SARS-CoV-2 vaccine in seronegative and seropositive participants in India"

Related to figure 2A

| | Overall cohort | |
|---|---|---|
| | Seronegative | Seropositive |
| **Covaxin** | | |
| Week 0 vs Week 4 | 0.0002 | 0.0037 |
| Week 0 vs Week 6 | <0.0001 | <0.0001 |
| Week 4 vs Week 6 | 0.0009 | 0.1280 |
| **Covishield** | | |
| Week 0 vs Week 4 | <0.0001 | <0.0001 |
| Week 0 vs Week 14 | <0.0001 | <0.0001 |
| Week 4 vs Week 14 | 0.0001 | <0.0001 |
| | **BBH** | |
| **Covaxin** | | |
| Week 0 vs Week 4 | 0.1283 | 0.1996 |
| Week 0 vs Week 6 | 0.0604 | <0.0001 |
| Week 4 vs Week 6 | 0.2133 | <0.0001 |
| **Covishield** | | |
| Week 0 vs Week 4 | 0.0007 | <0.0001 |
| Week 0 vs Week 14 | <0.0001 | <0.0001 |
| Week 4 vs Week 14 | 0.0002 | 0.0030 |
| | **KEM** | |
| **Covaxin** | | |
| Week 0 vs Week 4 | 0.0059 | <0.0001 |
| Week 0 vs Week 6 | 0.0015 | <0.0001 |
| Week 4 vs Week 6 | 0.0054 | 0.0022 |
| **Covishield** | | |
| Week 0 vs Week 4 | 0.0081 | <0.0001 |
| Week 0 vs Week 14 | <0.0001 | <0.0001 |
| Week 4 vs Week 14 | 0.3420 | <0.0001 |
| | **SUHRC** | |
| **Covaxin** | | |
| Week 0 vs Week 4 | 0.3932 | 0.0101 |
| Week 0 vs Week 6 | 0.0004 | <0.0001 |
| Week 4 vs Week 6 | 0.0018 | 0.0012 |
| **Covishield** | | |
| Week 0 vs Week 4 | 0.0051 | <0.0001 |
| Week 0 vs Week 14 | <0.0001 | <0.0001 |
| Week 4 vs Week 14 | 0.9652 | 0.1407 |
| | **SJRI** | |
| **Covaxin** | | |
| Week 0 vs Week 4 | 0.0780 | 0.2281 |
| Week 0 vs Week 6 | 0.0696 | 0.0565 |
| Week 4 vs Week 6 | 0.5751 | 0.6767 |
| **Covishield** | | |
| Week 0 vs Week 4 | 0.0117 | <0.0001 |
| Week 0 vs Week 14 | <0.0001 | <0.0001 |
| Week 4 vs Week 14 | 0.1217 | 0.0181 |

**Supplementary table 2 Change in antibody titers over time within each study group in each cohort** Data are p-values of two-way ANOVA with Tukey’s correction to compare differences over time in each cohort within each study group.

Related to figure 2B

Supplementary Figure 1A)

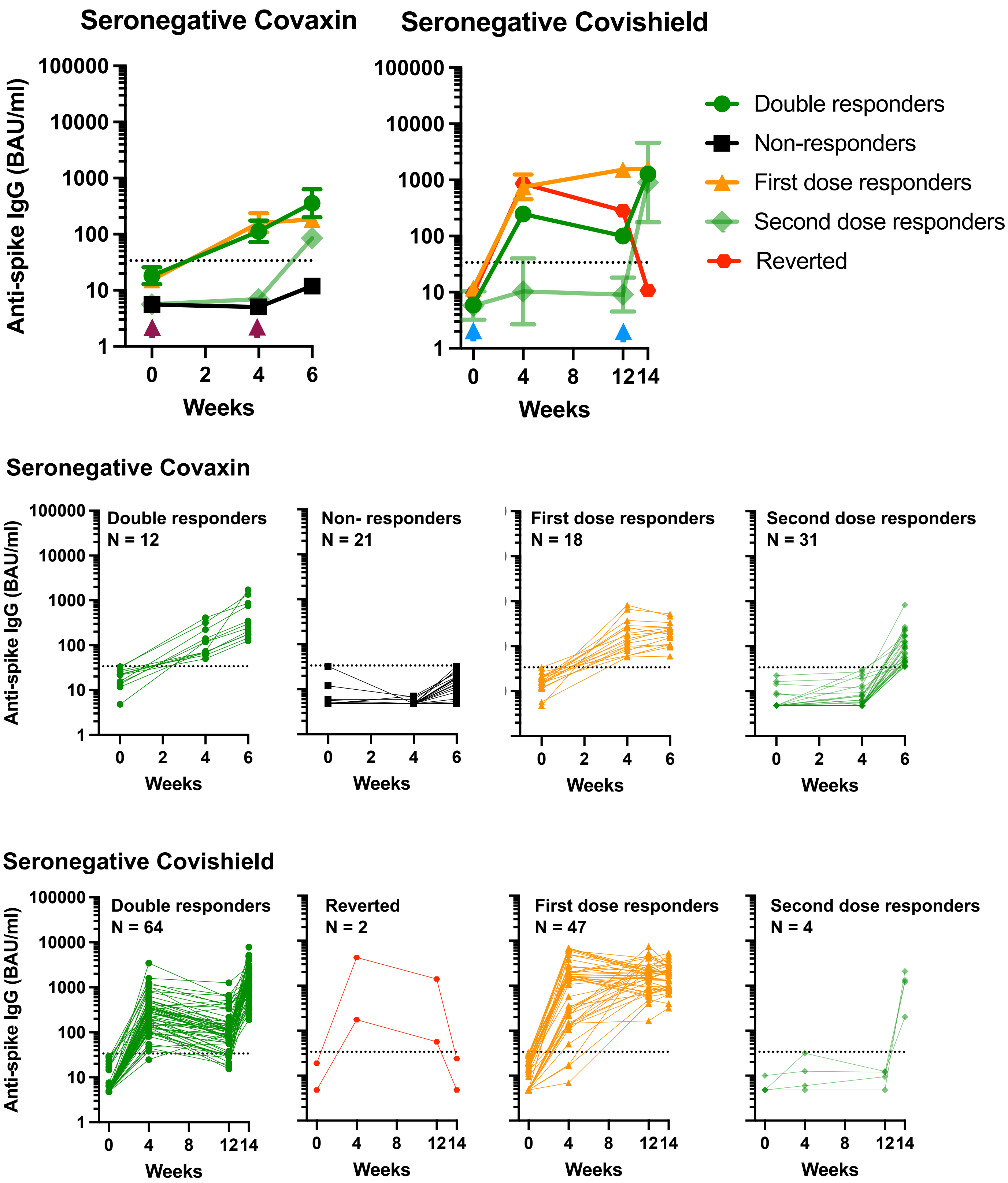

Supplementary Figure 1B)

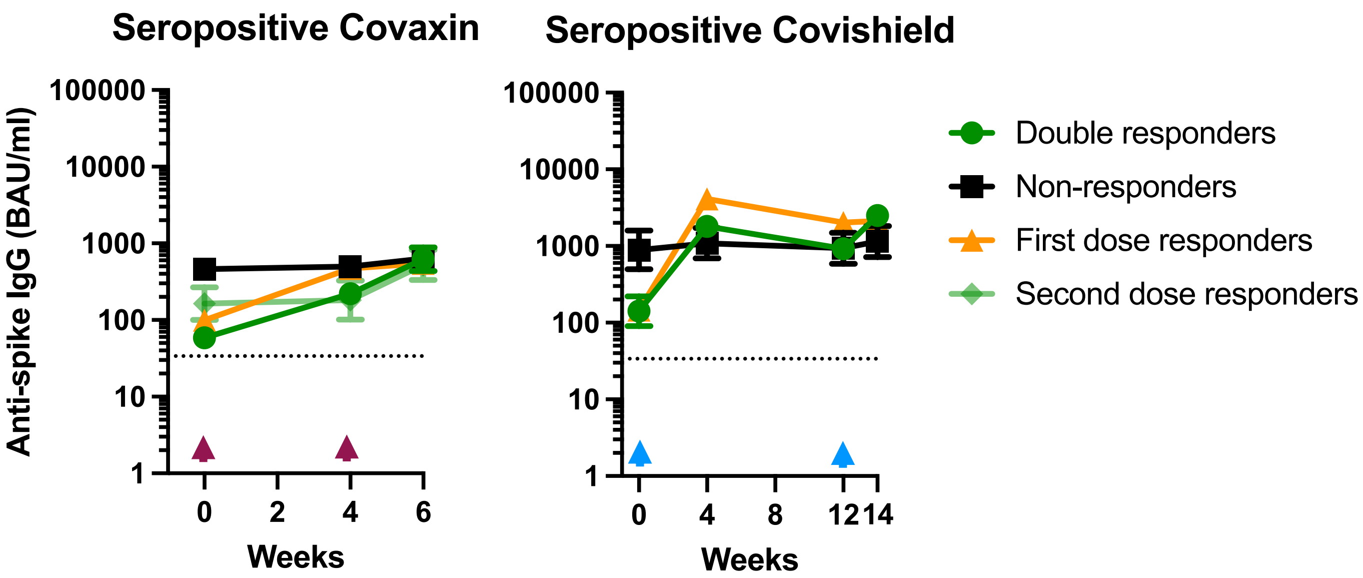


Seropositive Covaxin

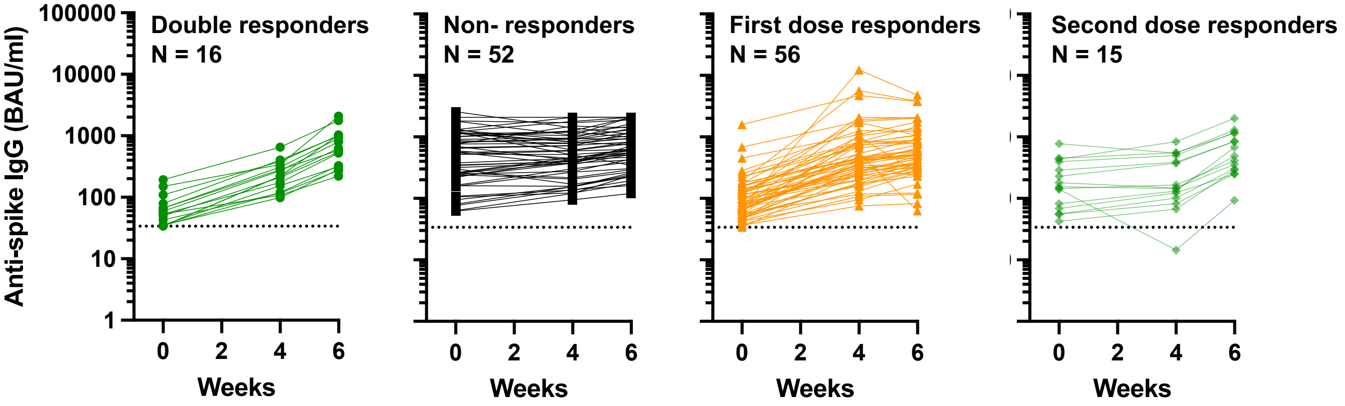


Seropositive Covishield

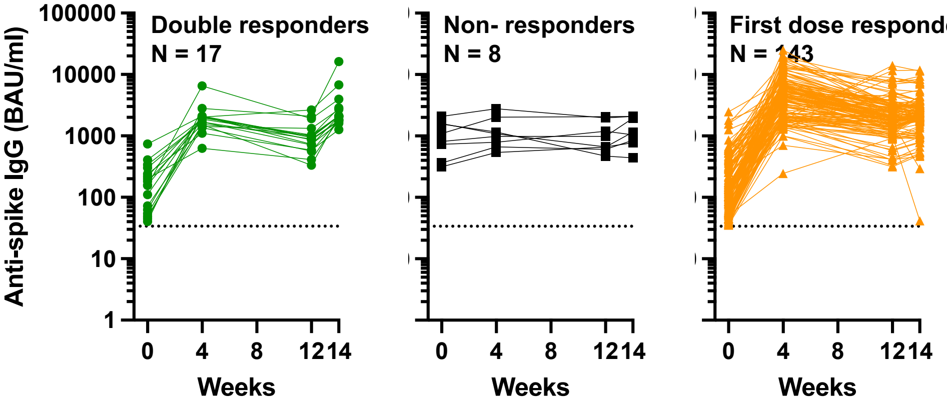


**Supplementary figure 1 Classification of study participants based on antibody trajectories following vaccination** A) Seronegatives B) Seropositives. Arrowheads indicate the time of vaccination

A)

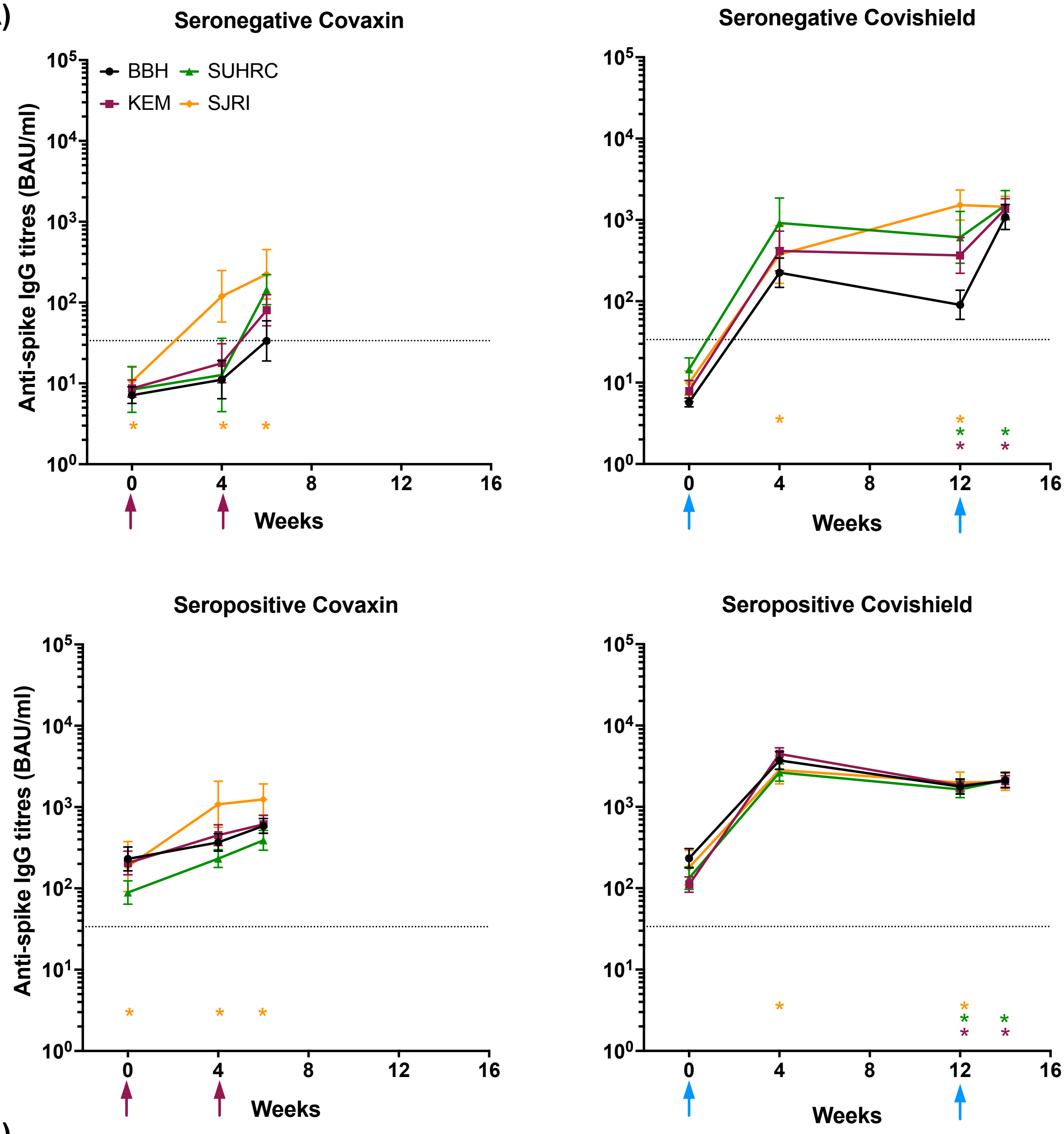


B)

| Covaxin | Seronegative Week 0 | Week 4 | Week 6 | Week 12 | Week 14 | Seropositive Week 0 | Week 4 | Week 6 | Week 12 | Week 14 |
|---|---|---|---|---|---|---|---|---|---|---|
| BBH vs. KEM | 0.4247 | 0.4135 | 0.4141 | | | 0.9926 | 0.5642 | 0.8504 | | |
| BBH vs. SUHRC | 0.8249 | >0.9999 | 0.3151 | | | 0.0476 | 0.0422 | 0.1298 | | |
| BBH vs. SJRI | 0.4136 | 0.1881 | 0.2646 | | | >0.9999 | 0.3516 | 0.1421 | | |
| KEM vs. SUHRC | 0.9996 | 0.5519 | 0.9914 | | | 0.0540 | 0.0022 | 0.0276 | | |
| KEM vs. SJRI | 0.9740 | 0.3481 | 0.5631 | | | 0.9987 | 0.4378 | 0.2217 | | |
| SUHRC vs. SJRI | 0.9980 | 0.1932 | 0.4845 | | | 0.6186 | 0.2486 | 0.0459 | | |
| **Covishield** | | | | | | | | | | |
| BBH vs. KEM | 0.1510 | 0.4588 | | 0.0501 | 0.9941 | 0.0036 | 0.9985 | | 0.8878 | 0.8025 |
| BBH vs. SUHRC | 0.0002 | 0.1040 | | 0.0164 | 0.8031 | 0.0199 | 0.0706 | | 0.6310 | 0.9317 |
| BBH vs. SJRI | 0.0381 | 0.5972 | | 0.0011 | 0.9955 | >0.9999 | 0.4477 | | 0.9925 | 0.8344 |
| KEM vs. SUHRC | 0.1467 | 0.6617 | | 0.4613 | 0.9027 | 0.8693 | 0.0235 | | 0.8948 | 0.9935 |
| KEM vs. SJRI | 0.9203 | 0.9934 | | 0.0181 | >0.9999 | 0.2299 | 0.3750 | | 0.8022 | >0.9999 |
| SUHRC vs. SJRI | 0.4730 | 0.5103 | | 0.2356 | 0.8804 | 0.3486 | 0.7973 | | 0.5703 | 0.9923 |

**Supplementary Figure 2. Change in antibody titers within each study group across cohorts**. Antibody responses against ancestral spike measured on the Diasorin TrimericS platform for per-protocol population across each site. Participants were classified as either seronegative or seropositive at baseline using the assay cut-off of 33.8 BAU/ml (horizontal dotted line). Stars within each graph indicate timepoints where sampling in greater than 25% of the population from the cohort was performed during the Omicron wave in January and February 2022. Colors of the stars indicate the cohort. Statistical comparisons between cohorts at each timepoint were performed using a two-way ANOVA with Tukey's correction. B) summary of p-values

Related to figure 2D

### Supplementary Figure 3A)

Uninfected cells    Infected cells (0.01 MOI)

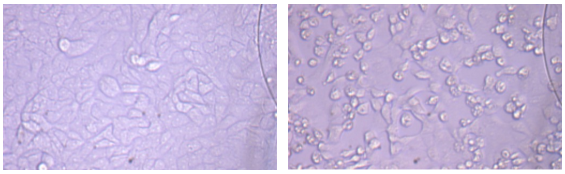

Infected cells + test sample at indicated dilution

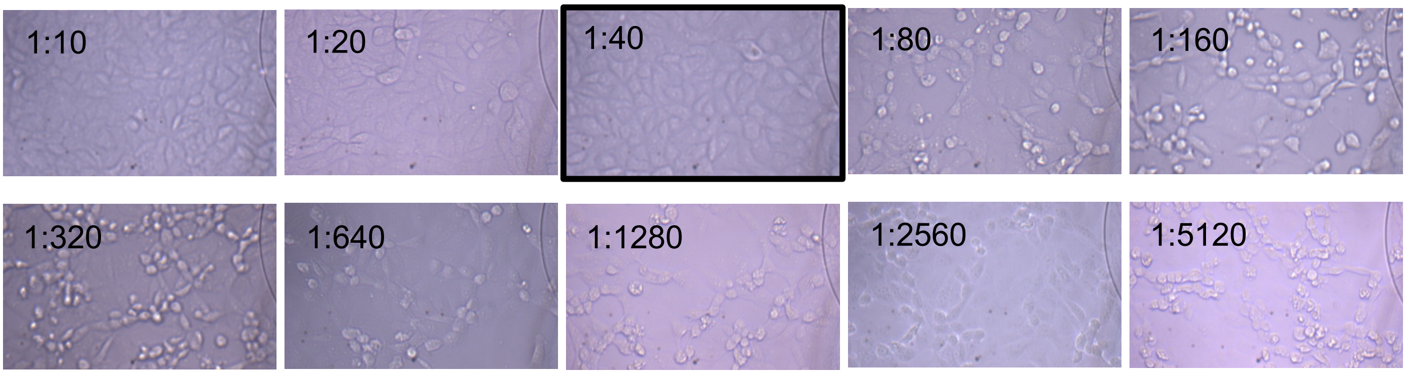


### Supplementary Figure 3B)

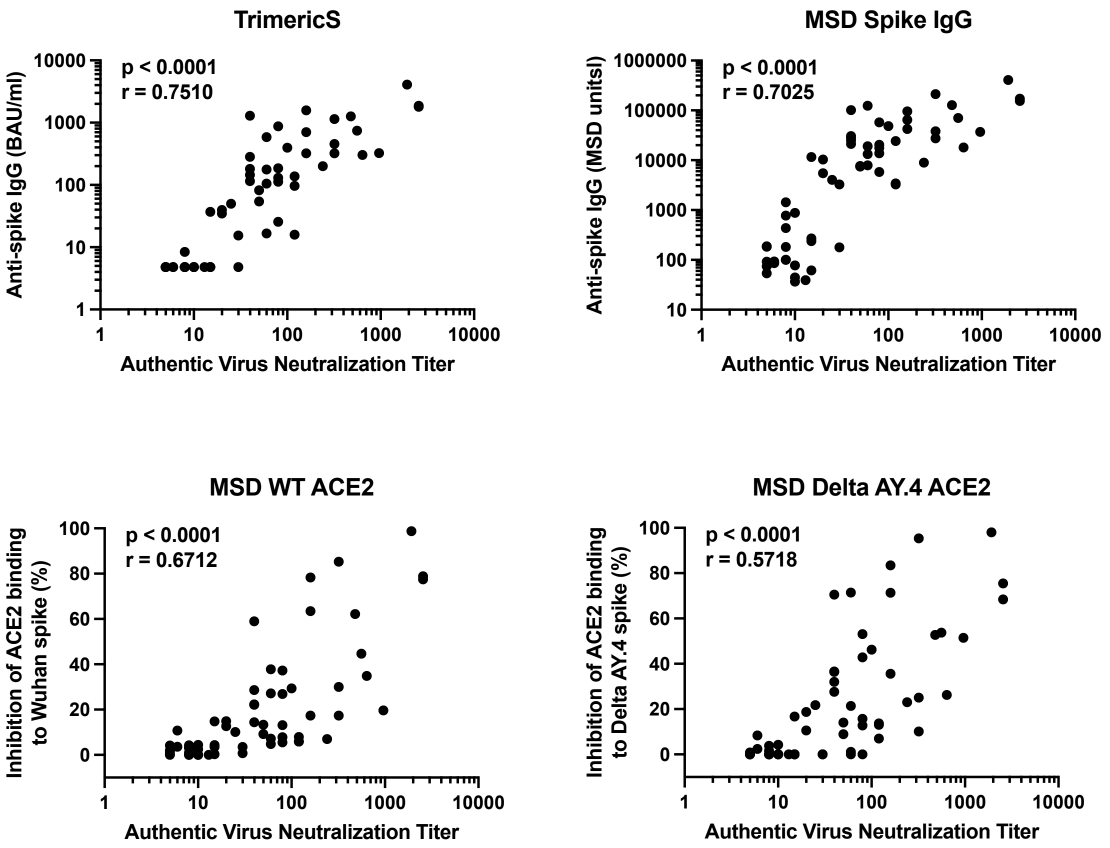

**Supplementary Figure 3C)**

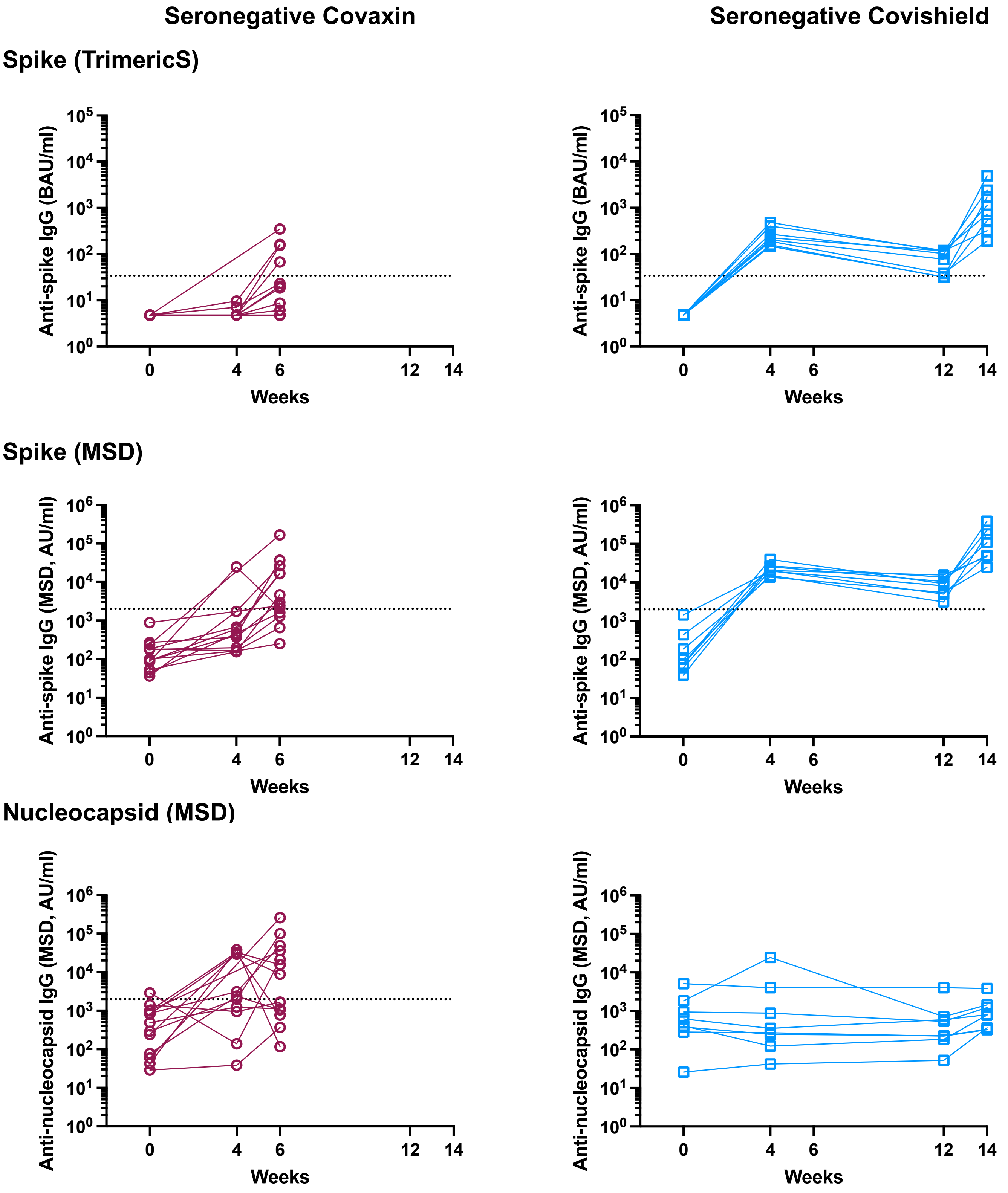

Supplementary Figure 3D)

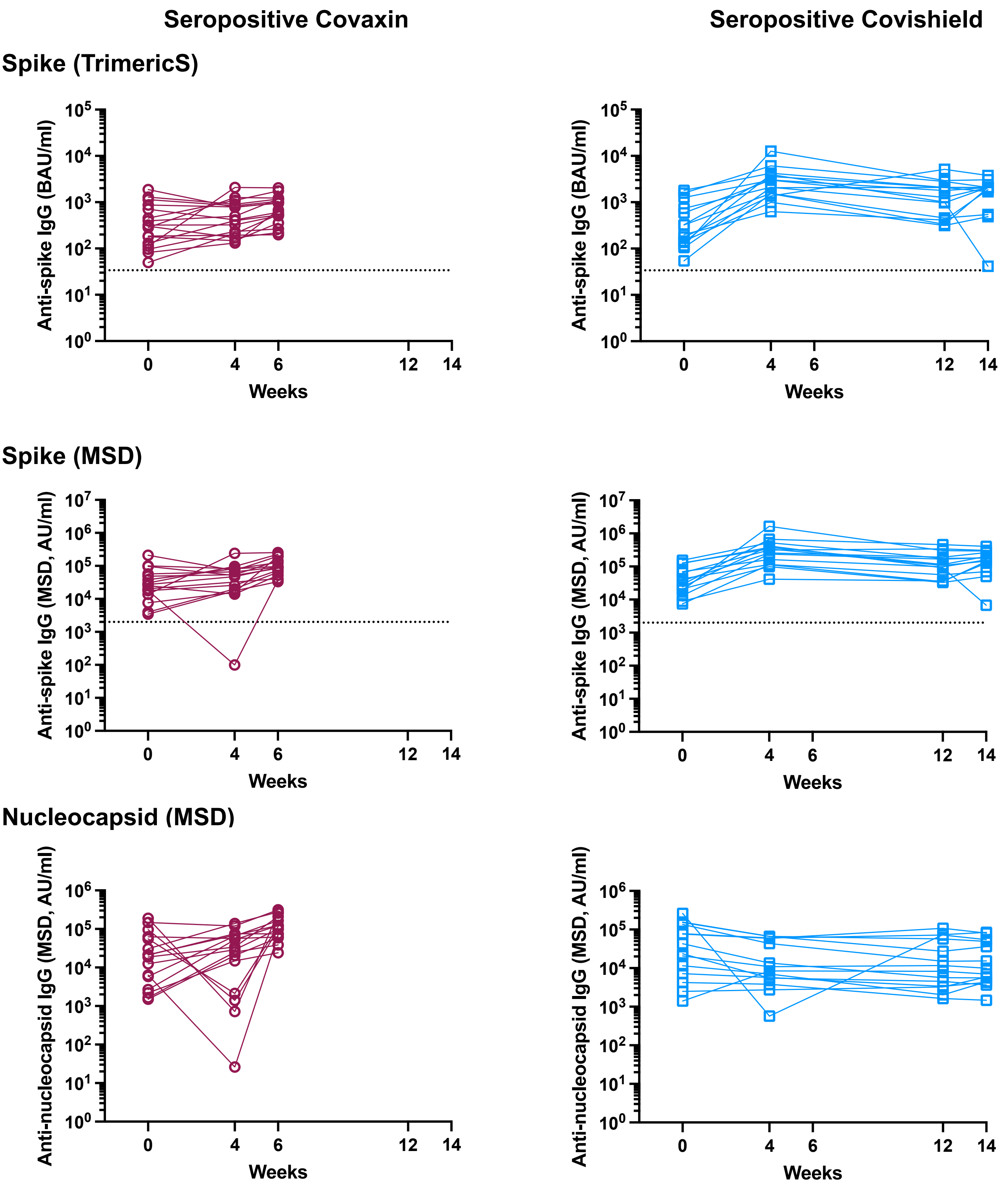


**Supplementary figure 3** A) Representative light microscope images showing determination of neutralizing titers. The highlighted image (1:40) shows the highest dilution of plasma at which there is complete rescue of virus-induced cytopathic effects. B) Pearson correlation between neutralization titers against authentic virus and either anti-spike binding IgG or percent inhibition of ACE-2 binding to ancestral spike protein. C,D) Binding antibody titers in participants who were included for neutralization assay from seronegative and seropositive groups respectively.

**Supplementary Figure 4A)**

Related to figure 3

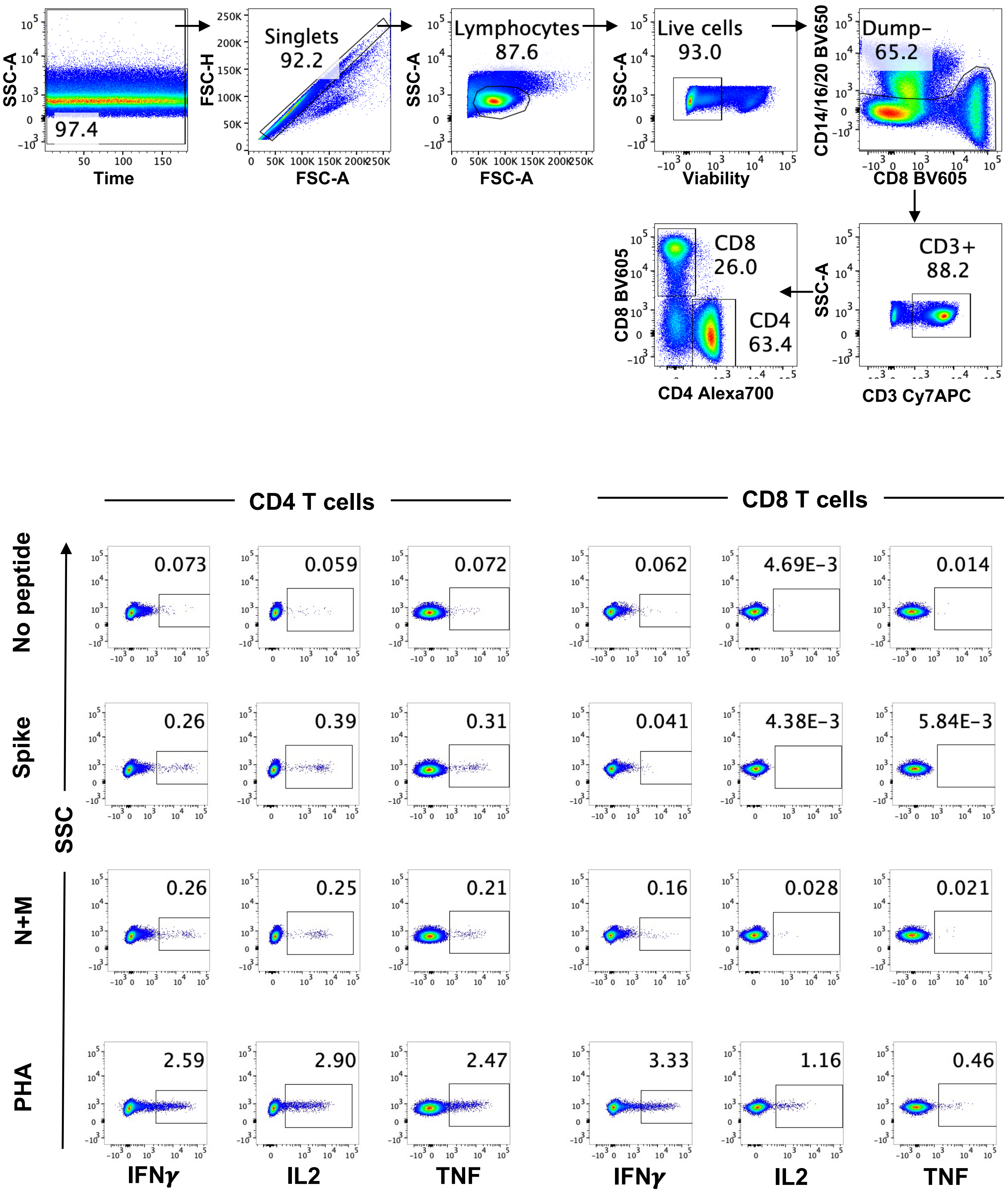

### Supplementary Figure 4B)

Related to figure 3A

**Seronegative Covaxin** | **Seronegative Covishield**

#### Spike (TrimericS)

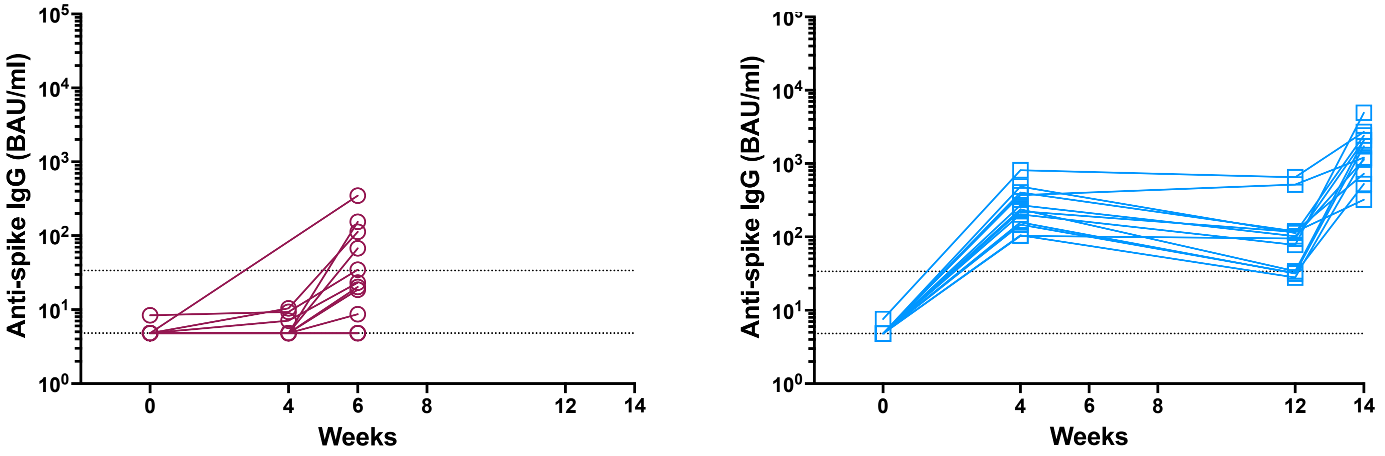


#### Spike (MSD)

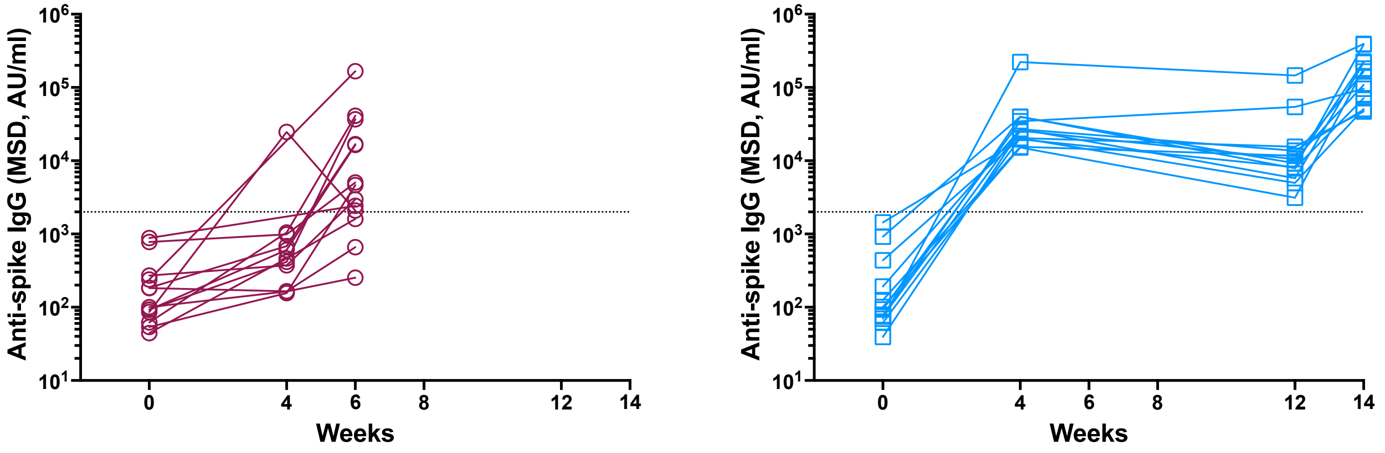


#### Nucleocapsid (MSD)

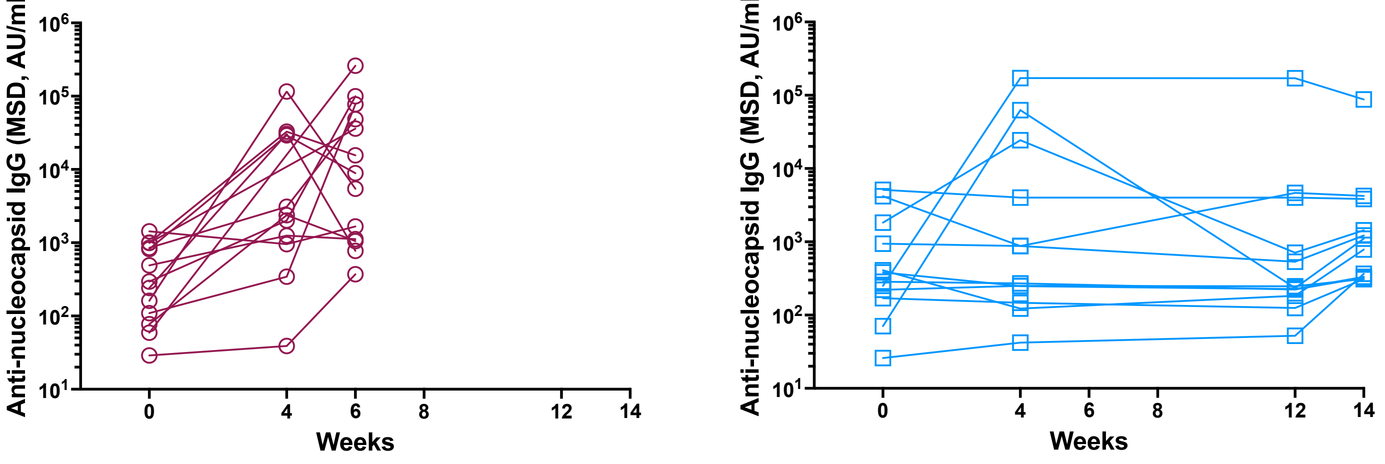



10

### Supplementary Figure 4C)

Related to figure 3B

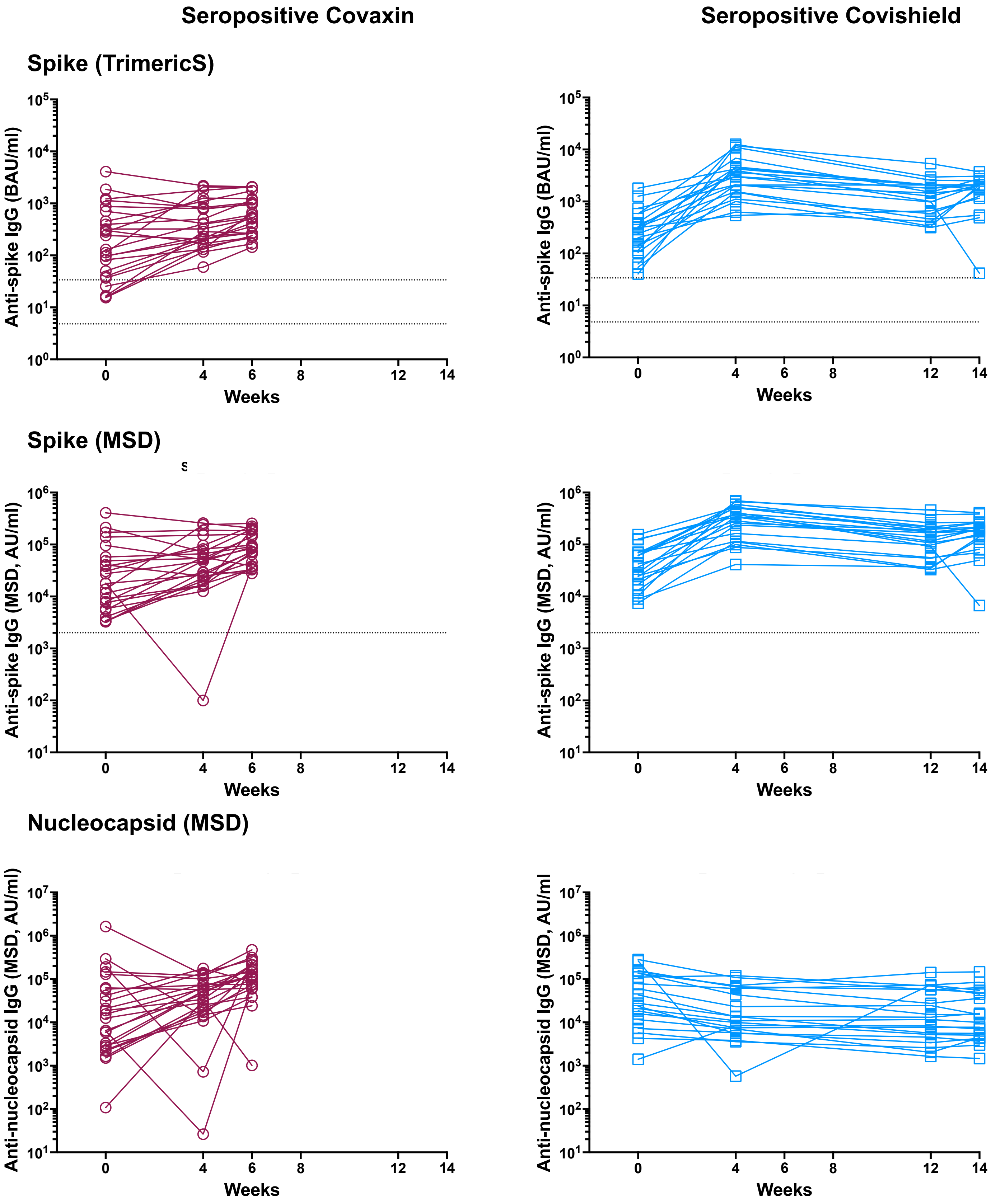


**Supplementary figure 4** A) Representative gating strategy for flow cytometry-based quantification of antigen-specific T cells expressing IFN$\gamma$, IL2 and/or TNF. B,C) Antibody responses against spike and nucleocapsid proteins in the seronegative and seropositive participants included in the ICS assay.

**Supplementary Figure 5A)**

Related to figure 3

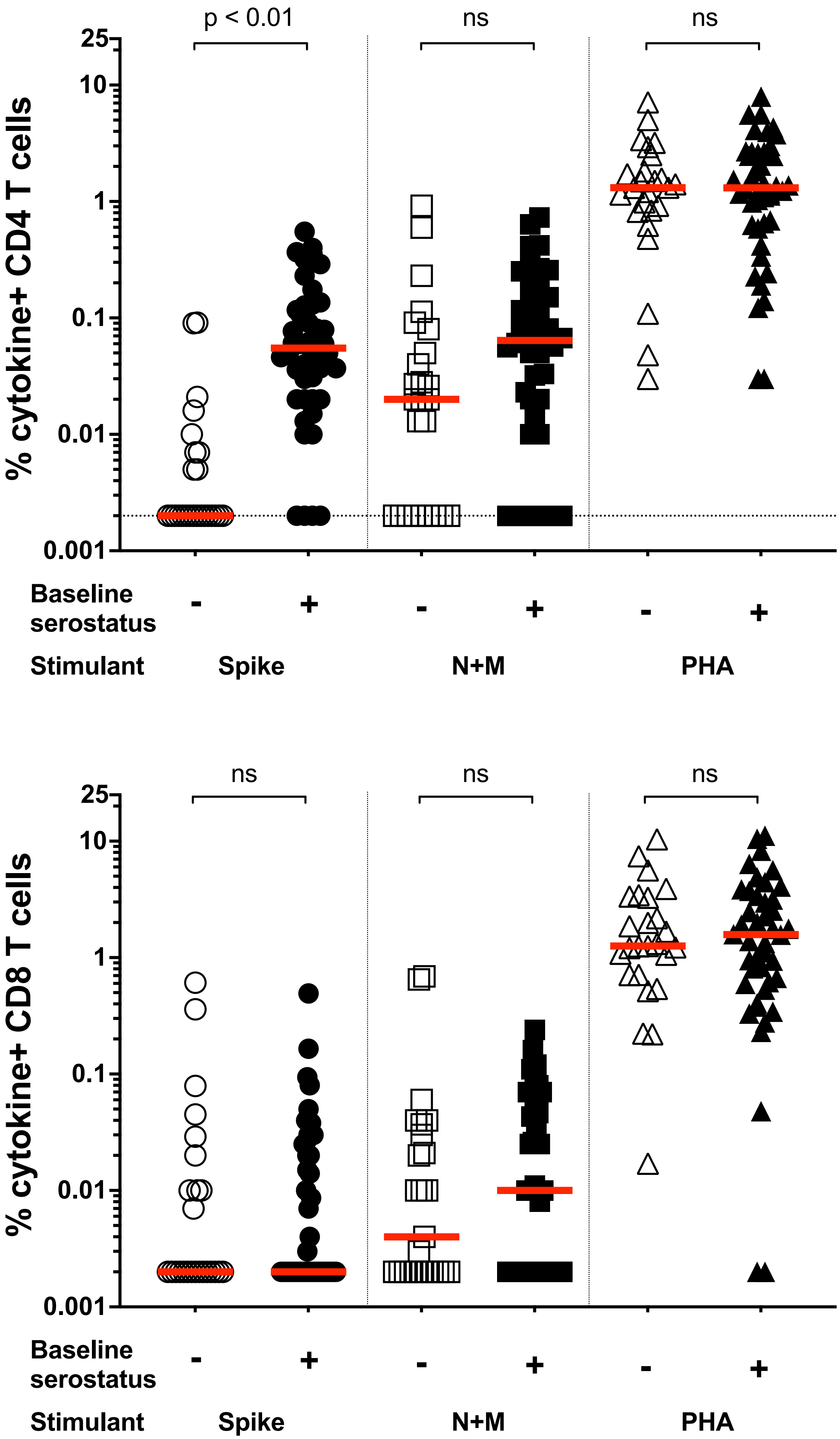

**Supplementary Figure 5B)**

Related to figure 3

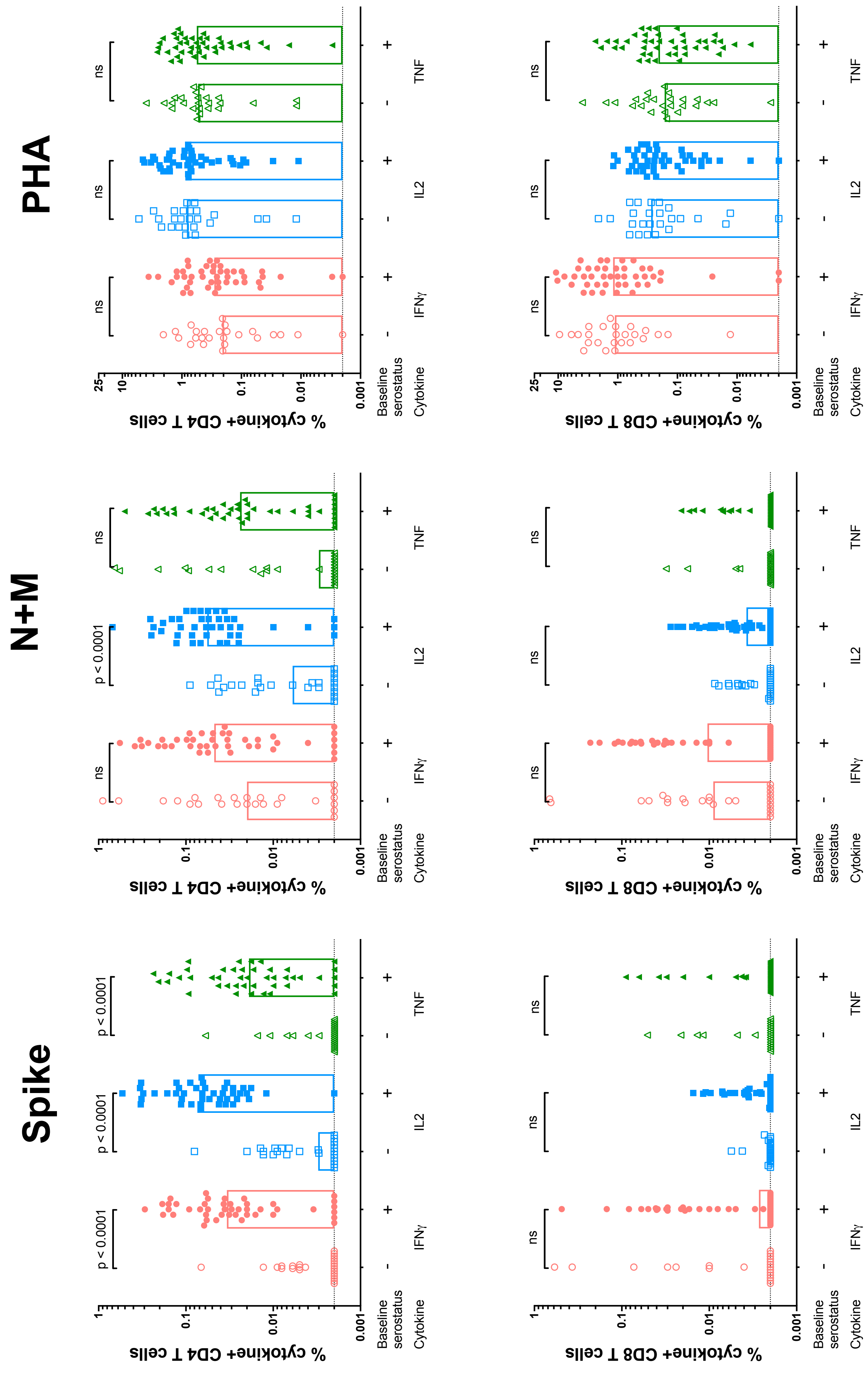

**Supplementary Figure 5C)**

Related to figure 3

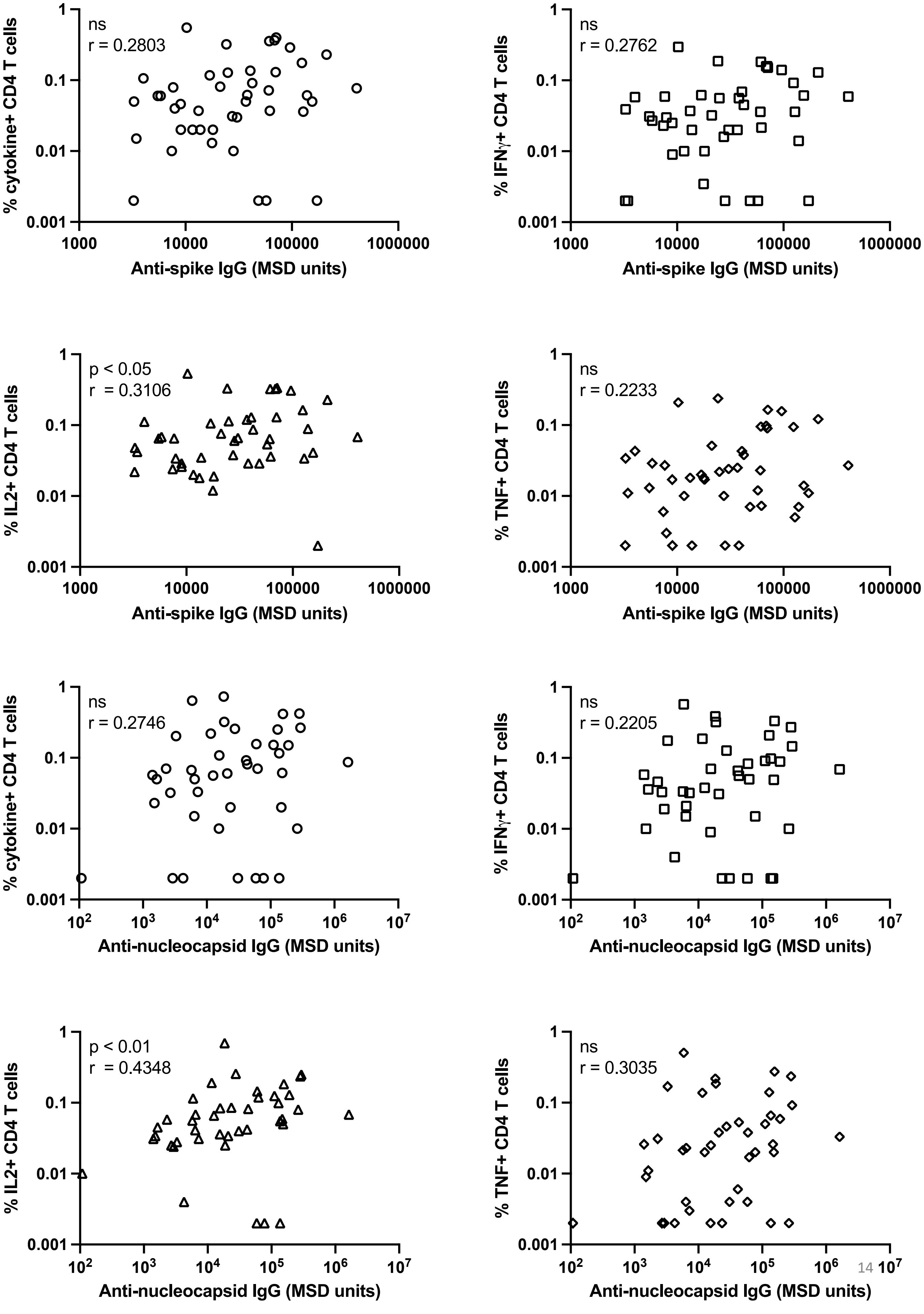

Related to figure 3

**Supplementary Figure 5 T cell responses prior to vaccination in seronegative versus seropositive participants.** A) T cell responses against a pool of peptides derived from either ancestral spike or a mixture N and M or control PHA measured by ICS in one of the cohorts. Background responses from an unstimulated control were subtracted. Participants were classified as either seronegative (n=25) or seropositive (n=44) against SARS-CoV-2 spike at baseline using the MSD assay. Solid horizontal line (red) indicates the group median. Differences between seronegatives and seropositives were assessed using Kruskal-Wallis test with Dunn's correction. B) Antigen-specific responses are shown for each of the indicated cytokines. Bar heights represent the group median. Differences between seronegatives and seropositives were assessed using Kruskal-Wallis test with Dunn's correction. C) Pearson correlation between antibody titers and spike-specific or nucleocapsid-specific CD4 T cells prior to vaccination

Related to figure 3A

Supplementary figure 6A)

TrimericS-based classification

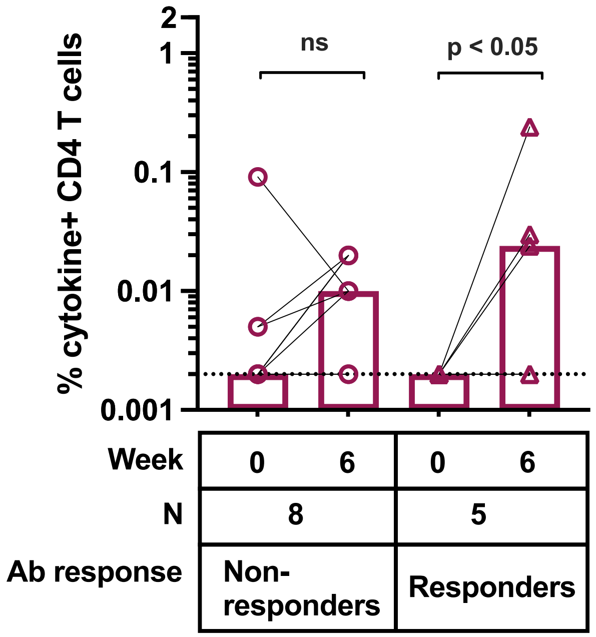


Supplementary figure 6B)

MSD-based classification

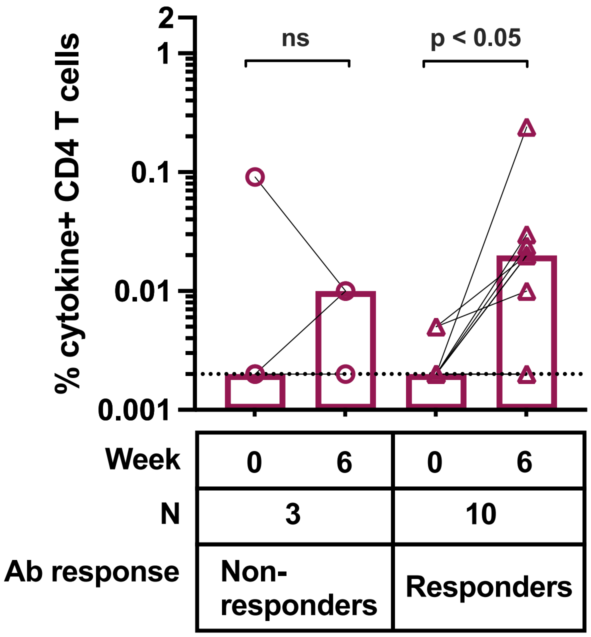

Supplementary figure 6C)

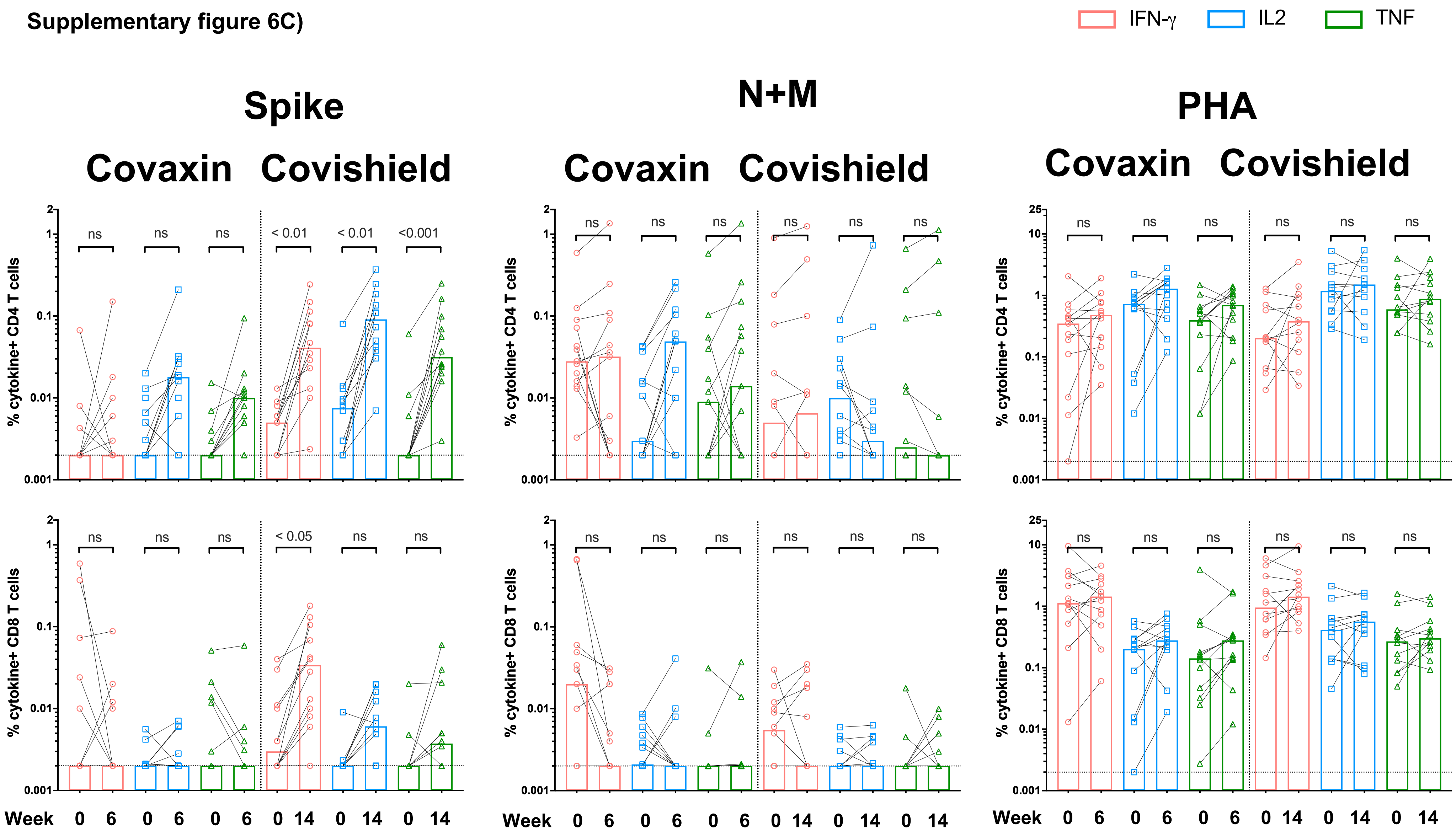


Related to figure 3A

**Supplementary Figure 6** A, B) Spike-specific T-cell responses among participants classified as either responders or non-responders based on antibody titers measured on A) TrimericS or B) MSD platform. Responders were classified as those whose post-vaccination antibody titers were greater than either 33.8 BAU/ml on Trimeric S or 1960 AU/ml (MSD units). Statistical comparisons were made using Kruskal-Wallis test with Dunn's correction. Bar heights represent group median

C) T-cell responses shown against individual cytokines pre- and post-vaccination against the indicated antigens (either spike, mixture of N and M or PHA). Statistical comparisons were made using Kruskal-Wallis test with Dunn's correction. Bar heights represent group median

Related to figure 3B

Supplementary figure 7A)

TrimericS-based classification

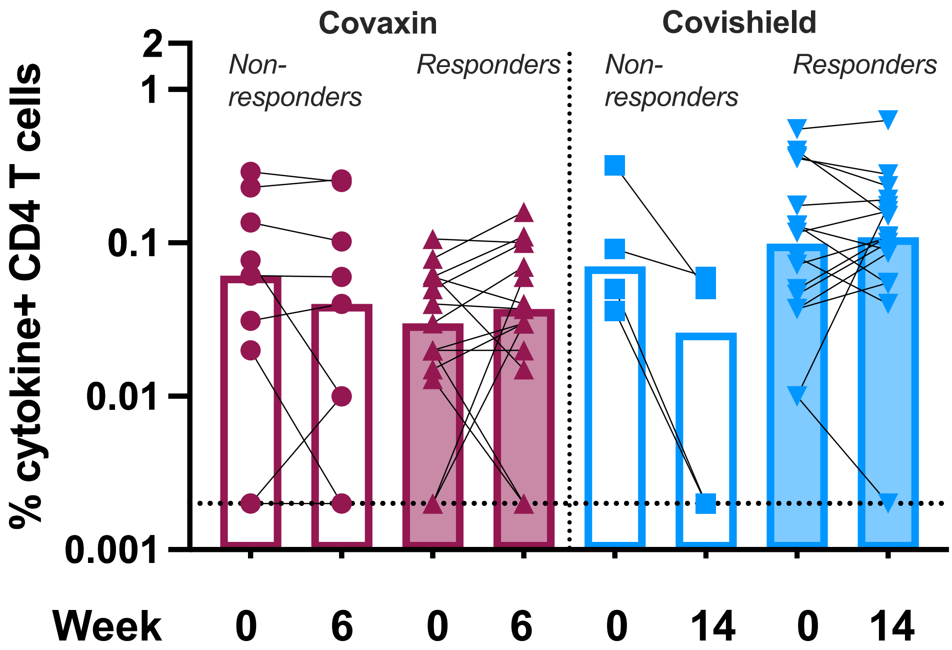


Supplementary figure 7B)

MSD-based classification

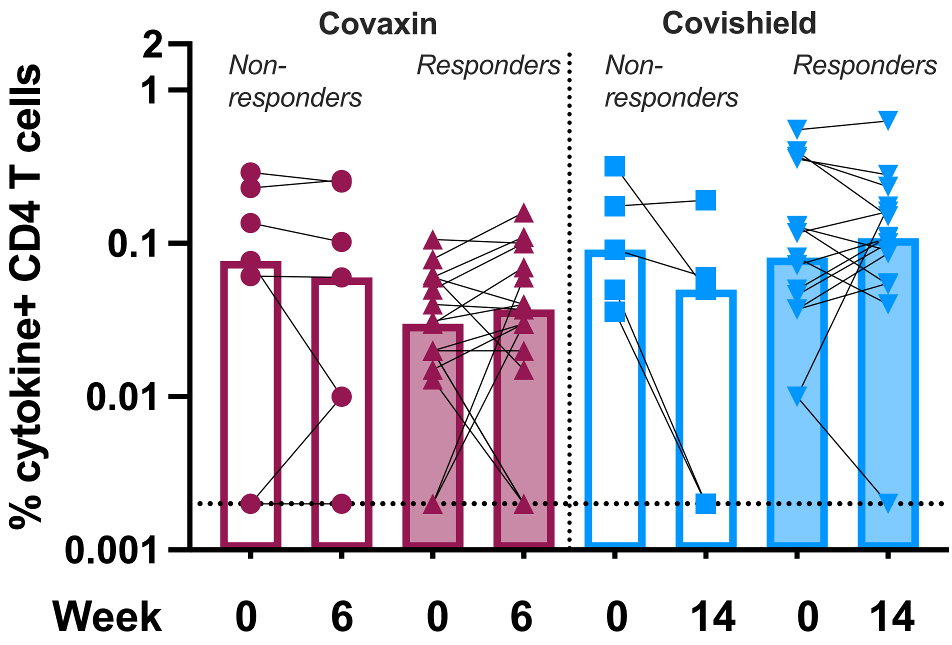

Related to figure 3B

**Supplementary figure 7C)**

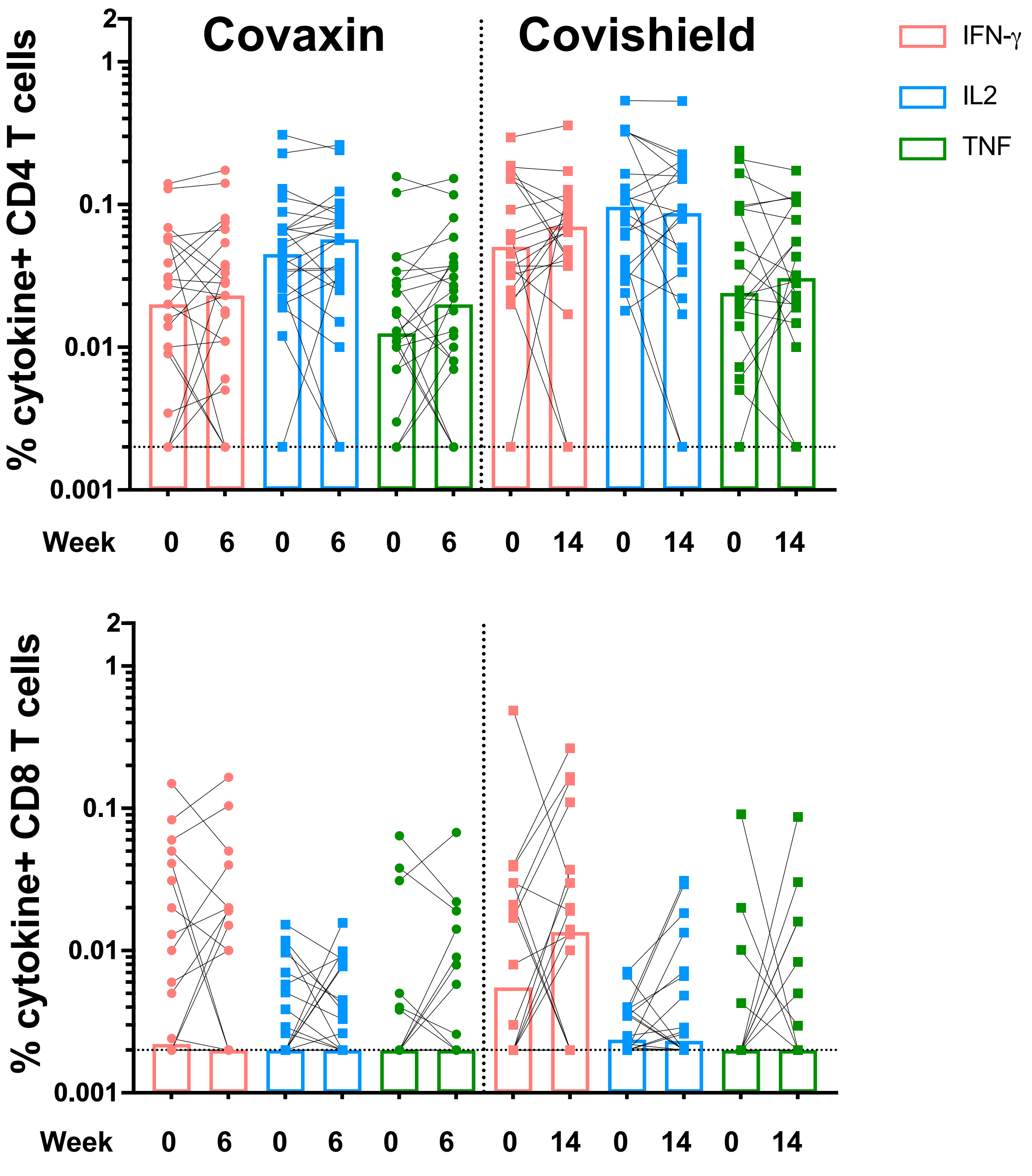


**Supplementary Figure 7**
A, B) Spike-specific T-cell responses among participants classified as either responders or non-responders based on antibody titers measured on A) TrimericS or B) MSD platform. Responders were classified as those whose post-vaccination antibody titers were atleast 2-fold greater than at baseline. Statistical comparisons were made using Kruskal-Wallis test with Dunn's correction. Bar heights represent group median
C) T-cell responses shown against individual cytokines pre- and post-vaccination against the indicated antigens (either spike, mixture of N and M or PHA). Statistical comparisons were made using Kruskal-Wallis test with Dunn's correction. Bar heights represent group median
